# A Randomised Controlled Trial of the effects of Galacto-Oligosaccharides on the gut brain-axis of young females

**DOI:** 10.1101/2025.01.09.25320270

**Authors:** Nicola Johnstone, Kathrin Cohen Kadosh

## Abstract

**Background:** Galacto-oligosaccharides (GOS) are prebiotics linked to gut microbiota modulation and potential gut-brain axis effects on neurochemistry, mental health and cognition. This study evaluated the influence of GOS along the gut-brain axis, integrating assessments of mental health, neurochemistry, gut microbiome composition, cognition, and nutrition in healthy females.

**Methods:** In a randomised, double-blind, placebo-controlled trial, 83 females (17–25 years) received GOS or placebo for 28 days. Assessments occurred at baseline, endline, and 28 days post-supplementation. The primary outcome was trait anxiety, with secondary outcomes including neurochemical measures of GABA and glutamate via 1H-MRS in the dorsolateral prefrontal cortex (dlPFC), anterior cingulate cortex (ACC), and inferior occipital gyrus (IOG) of the brain, and gut microbiome composition. Tertiary outcomes included social anxiety, depression, emotion behaviour, reaction times, and nutritional intake. Analyses included intention-to-treat, per-protocol, and sensitivity approaches.

**Results:** Trait anxiety did not differ significantly between groups at endline (p = .443), though trends favoured lower anxiety in the GOS group at follow-up (p = .069). GOS reduced GABA at trend significance in the inferior occipital gyrus (p = .053) and dorsolateral prefrontal cortex (p = .088) in high-anxious participants, with effects persisting at follow-up. GOS increased Bifidobacterium abundance (p = .001) but did not affect overall microbiome diversity. Tertiary outcomes showed no significant changes in social anxiety or depression but faster reaction rates in high-anxious participants for simple (p = .036) and choice tasks (p < .001). Nutritional intake was unaffected.

**Conclusion:** While GOS supplementation did not significantly reduce trait anxiety, it produced neurochemical changes and transient modulations of the gut microbiome in Bifidobacterium abundance. These findings suggest GOS-induced changes can be traced along the gut-brain axis, with implications for mental health and cognitive function that warrant further investigation.

## Introduction

Mental health challenges in youth have long-lasting consequences, impacting both individuals and society as a whole. ^1,2^. Novel interventions for youth mental health are a priority, especially in cases where pharmacological therapies may be less appropriate. One approach is to modify the gut microbiome to guide holobionts towards a homeostatic state with an enhanced capacity to thrive. Over the past 15 years, extensive research has highlighted the inextricable links between the gut microbiota and the brain, offering a promising pathway for intervention. However, deterministic research in humans remains limited.

The gut microbiome is an entity engaged in complex interactions in human homeostasis ^3^, following a developmental trajectory through successive stages to reach relative stability in adulthood ^4^. That said, gut microbiota can be adjusted for health benefits through environmental or dietary means in a matter of days ^5^, although effects appear transient ^6^. Of gut microbiome modifiers, prebiotics are most extensively studied. Thirty years ago, Gibson and Roberfroid introduced the term prebiotics, to describe the gut health benefits of non-digestible dietary fibres^7^. Advances in methodology have since broadened this concept to include systemic health benefits beyond the gut ^8,9^, and the use of prebiotics to modulate the gut microbiome has proved revealing of the inextricable links between the gut, the brain, and behaviour.

Beneficially modifying the gut microbiome with prebiotics has shown anxiolytic effects in high-anxious females ^10^ , demonstrating that increasing *Bifidobacterium* abundance through prebiotic supplementation can significantly alleviate anxiety symptoms. Extensive evidence from preclinical models reveals the crucial role of the gut microbiome in regulating brain function and emotional behavior ^12–15^. During development, neurobiological systems are fine-tuned, setting maturational trajectories. As the brain grows and adapts, the gut microbiota also changes ^16–18^, making it a promising modifiable target for guiding neurodevelopment ^19,20^. In addition, preclinical research shows that the gut microbiome plays a crucial role in mental health disorder emergence ^21–24^. While the mechanisms of how this occurs is not yet clear, influential neurotransmitters may be one potential pathway.

When the composition and activity of gut bacteria are adjusted, the production and regulation of key neurotransmitters, such as gamma-aminobutyric acid (GABA) are affected, thereby influencing brain function and mental health outcomes. The gut microbiome may contribute to GABA levels in the brain via various processes. For example, several species of *Bifidobacteria*, (*B. adolescentis, B. dentium* and *B, infantis*) are known to produce GABA in vivo in the gut ^25,26^ that may contribute to circulating peripheral GABA levels ^27^. Another way may be via short-chain fatty acids produced in the gut (e.g., butyrate, propionate and acetate) that serve as precursors of neurotransmitters ^28,29^. While neither of these are direct effects, mechanistic actions are likely facilitated by humoral or neuronal pathways ^30,31^. However, precise mechanisms of the effect of the gut microbiota on the brain are still debated.

GABA and glutamate are the primary inhibitory and excitatory neurotransmitters of the brain, essential for synapse formation and functional connectivity. Early brain development relies on GABA-triggered glutamate responses that shape neurocircuitry ^32,33^. The developmental trajectories of glutamate and GABA measured by magnetic resonance spectroscopy in humans, are non-linear and interconnected ^34^. Glutamate levels steadily decline before stabilising in adulthood, whereas GABA levels decrease during early childhood, increase during adolescence, and stabilise thereafter ^34–36^. This results in a net reduction in the glutamate-to-GABA ratio with age, reflecting a progressive balance in the excitatory/inhibitory (E/I) ratio.

During adolescence sensitive periods marked by changes in the E/I ratio support neuroplasticity critical for cognitive development ^37–40^. These neurochemical shifts coincide with the peak onset of mental health disorders, such as anxiety, emphasising the need to understand how fluctuations neurochemical balance relate to psychological maturation ^41^. Stable brain function depends on maintaining a balance between excitatory and inhibitory neural circuits ^42,43^ with perturbations resulting in the emergence of divergent states, shifting the brain towards sensory-driven (bottom-up) dominance over cognitive (top-down) control^43^. This imbalance reflects insufficient inhibition and heightened excitability (glutamate activity), potentially contributing to mental health challenges. Restoring balance by enhancing GABA-mediated inhibition could stabilise neural activity and promote improved psychological well-being.

Supporting healthy GABA regulation through low-impact methods, such as cultivating a healthy gut microbiome, may help maintain brain health and psychological wellbeing. In a pioneering interventional trial using multi-level assessments of mental health, behaviour, gut microbiome, and dietary intake, we showed that the prebiotic galacto-oligosaccharide (GOS) has anxiolytic effects in highly anxious females aged 18-25 ^10^. Results illustrated that GOS improved self-reported anxiety levels, reduced attentional bias to negative stimuli, and promoted the growth of *Bifidobacterium*, altering the gut microbiome composition in highly anxious participants. There were further dietary changes associated with the increase in *Bifidobacterium* abundance whereby intakes of carbohydrates and sugars decreased and fat increased ^44^. In the current study, we aim to replicate these effects in addition to investigating how GOS impacts GABA and glutamate levels in the brain. We anticipate that GOS will elevate GABA levels in key brain regions integral to emotion regulation. These include the dorsolateral prefrontal cortex (dlPFC), a late maturing region weighted towards emotion regulation, and the anterior cingulate cortex (ACC) a region associated with higher order cognitive control ^45^. As a control, inferior occipital gyrus (IOG), an early maturing region weighted towards visual processing with limited direct input into emotion regulation processes will also be included.

A further 28 day follow up period will determine longevity of effects at each level of assessment. It is predicted that participants receiving GOS will, in comparison to the placebo group;

(i) In the primary outcome, reduce trait anxiety levels.
(ii) In the secondary outcomes, increased levels of GABA in specific brain regions involved in emotional regulation such as the dorsolateral prefrontal cortex (DLPFC) and the anterior cingulate cortex (ACC), and increase *Bifidobacterium* abundance in the gut microbiota.
(iii) In the tertiary outcomes, exhibit improvements on other indices of mental health, and elicit enhanced emotion behaviour and cognitive function, and alter nutritional intake consuming less carbohydrate and sugar, and increased fat.

## Method

This study obtained favourable ethical approval from the University ethics committee. Written informed consent was secured from all participants, with primary caregivers providing additional consent for those under 18 years old to participate in MRI scans. Participants provided assent before undergoing MRI procedures. Compensation of £75 was provided to participants for their time. The protocol for this study was registered on https://clinicaltrials.gov/ [NCT03835468] on February 2, 2019.

### Participants and design

This two-arm double-blind placebo-controlled trial aimed to recruit 83 females aged 17-25 between April 2022 and May 2023. Sample size was calculated for a medium sized effect on the primary outcome with 80% power and alpha level .05 for an ancova model with four covariates. Eligibility criteria included females aged 17-25 with no clinical anxiety, co-morbid psychological diagnoses, developmental disorders, gastrointestinal issues, restrictive diets, or MRI contraindications. Recruitment occurred through word of mouth and invitations extended to existing participant databases. The flow of participants is outlined in **Figure 1**.

**Figure 1.**
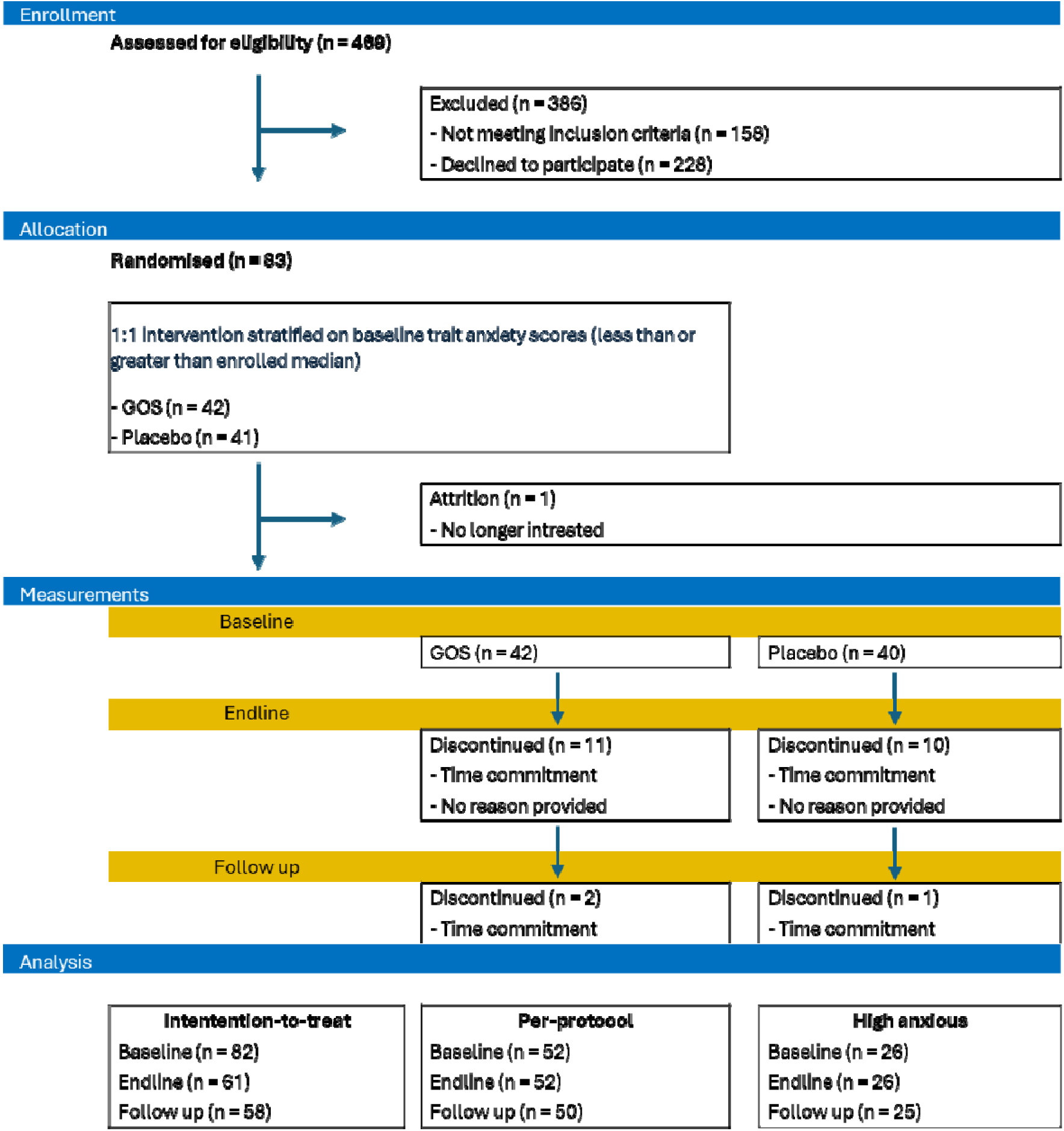
Participant flow chart showing the number of participants progressing through the study milestones.

Of 469 screened, 158 did not meet inclusion criteria and 228 declined to participate. Eighty-three females enrolled, with allocation to supplement intervention groups (1:1) based on stratification of trait anxiety scores to ensure equitable distribution of anxiety levels using custom software created by author NJ. The study research assistant enrolled and assigned participants using the software. 42 were allocated to the GOS group, and 40 to the placebo group. In total, 24 participants discontinued, 13 in GOS group, and 11 in placebo group.

Analyses were performed on three populations within the sample, (i) Intention-to-treat (ITT), which includes all randomised subjects regardless of protocol adherence or completion of primary measures at endline (T2) or follow-up (T3); (ii) Per-Protocol (PP), comprising subjects who completed the supplement intervention with at least 80% compliance and provided primary trait anxiety measures at baseline (T1) and endline (T2), considered complete cases; and (iii) a subgroup for sensitivity analysis, focusing on individuals with high trait anxiety, defined as those scoring above the sample median on the STAI trait subscale at T1, and analysed within PP populations. No adverse events were reported, and discontinuation was attributed to either time commitments or loss of contact.

Allocation concealment was maintained with the GOS and placebo supplements provided in powdered form in plain, matched, coded sachets by FrieslandCampina, Amersfoort, the Netherlands. These were 7.5 g/day (corresponding with 5.5 g/day active Biotis® GOS (FrieslandCampina, Amersfoort, the Netherlands) and 7.5 g/day Maltodextrin Maltosweet G 181 (Tate & Lyle) as placebo. Outcomes were assessed at baseline (T1), endline (T2), and 4 weeks later (follow-up, T3). Primary outcomes assessed trait anxiety using the state-trait anxiety inventory ^46^. Secondary outcomes measured neurochemical levels (GABA) using ^1^H-MRS and gut microbiome composition via metagenomic shotgun sequencing. Tertiary outcomes included social anxiety (SAS ^47^), depression (BDI-II, ^48^), emotion behaviour (dot probe task ^49^), reaction time ^50^, nutritional intake, and longevity of effects post supplement intervention, at follow up.

### Procedure and materials

We adapted a previous RCT ^10^ by extending the study duration and incorporating MRI measurements (**Figure 2**). In brief, at each time point (T1, T2, T3), participants completed a battery of questionnaires hosted online ^51^ on trait anxiety ^46^, social anxiety ^47^, depression ^48^, sleep quality ^52^, menstrual symptoms ^53^, physical activity ^54^ and demographics, and kept a food diary in the 4 days prior to in-person testing. In-person testing involved a computer tasks (dot probe ^49^, reaction times ^50^) and MRI scans lasting approximately 1 hour. MRI scans comprised anatomical images for constructing volumes of interest (VOIs) for MR spectroscopy, as well as 3 ^1^H-MRS scans. For the ^1^H-MRS scans, three VOIs were defined over the left dorsolateral prefrontal cortex (dlPFC), anterior cingulate cortex (ACC), and the left inferior occipital gyrus (IOG). Stool sample collection kits were provided for stool collection at home for later gut microbiome analysis. At T1 only, participants received a 28- day supply of supplements, instructed to consume 1 sachet daily at breakfast, mixed in food or a drink and return empty sachets for compliance monitoring. Stool sample collection kits were provided for stool collection at home for later gut microbiome analysis.

**Figure 2.**
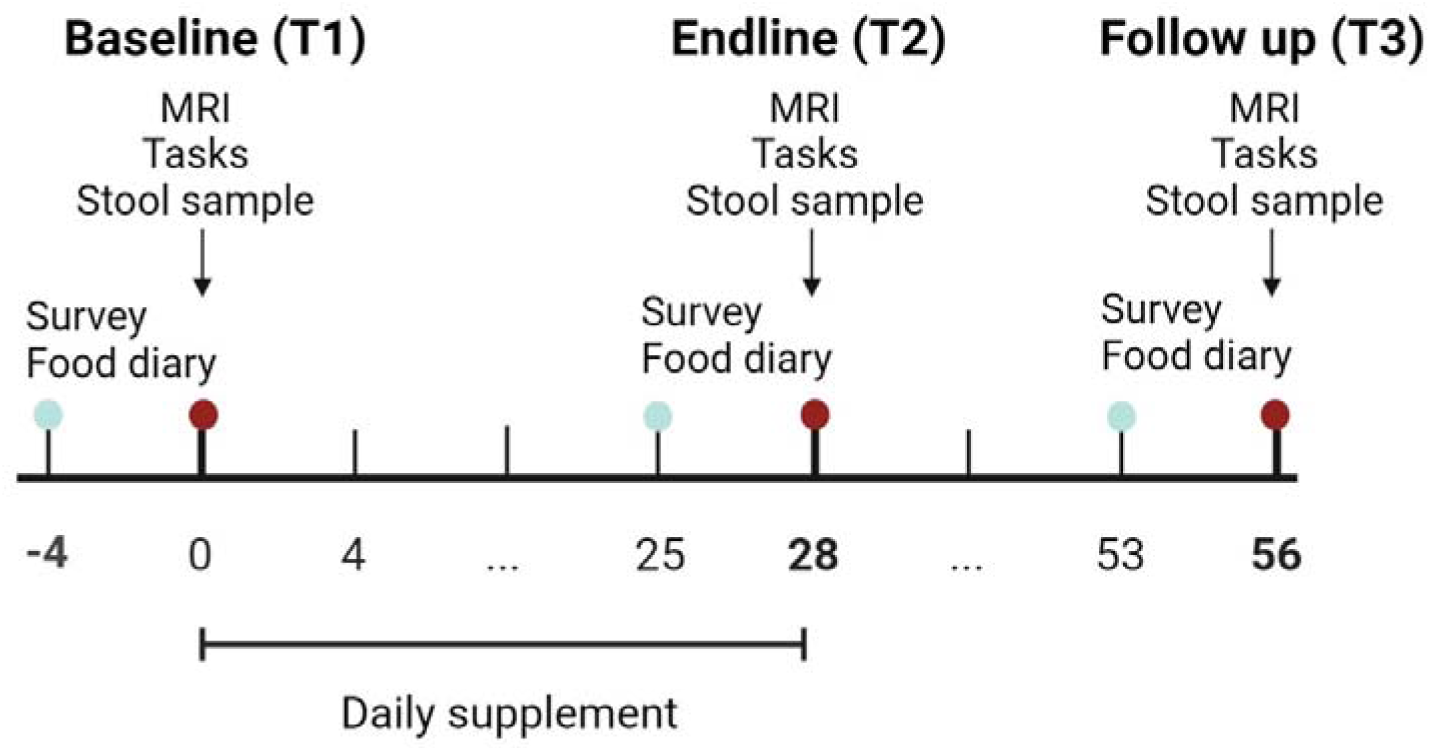
The study protocol illustrates the three testing points over 56 days. Day Zero represents baseline measurement (T1), followed by a 28-day supplement intervention. Endline measures (T2) were taken on the last day of supplement consumption, with a final follow-up session (T3) around day 56. The duration for recording food diaries is indicated. Survey measures were conducted using online platforms, including validated questionnaires for trait anxiety, depression, and demographic variables. In-person testing involved an MRI scan and completion of two behavioural tasks on a laptop outside the scanner. Stool sample collection kits were provided for participants to collect and return samples to the lab for storage, prior to microbiome analysis.

### Questionnaires

#### Demographic

measures collected by self-report included ethnicity, diet, height in centimetres and weight in kilograms (validated at in person sessions) and date of birth. Age was calculated from the reported date of birth and testing date. Data on hormonal contraceptive use and menstrual cycle phase was collected (average duration of menstrual cycle over the past 3 months, date of last period) and calculated in reference to testing date. Illness occurrence over the previous 4 weeks was also recorded.

#### Trait anxiety

The trait scale of the State-Trait Anxiety Inventory (STAI, ^46^), is a validated scale measuring anxiety over the previous 2 weeks, comprised of 20 questions assessing feelings related to trait anxiety in general. All items are rated on a 4-point (1-4) scale (e.g., from “Almost Never” to “Almost Always”), and summed to a total anxiety score with a range from 20-80. Higher scores indicate higher trait anxiety levels. Reliability across all assessments was good (20 items α = .918).

#### Social anxiety

The Social Anxiety Scale for Children-Revised (SAS, ^47^) is a 22 item validated self-report measure of 3 factors related to social anxiety; fear of negative evaluation (8 items); social avoidance and distress in new situations (6 items) and social avoidance and distress in general (4 items). Items are rated on a 5-point Likert scale (1 = not at all, 2 = hardly ever, 3 = sometimes, 4 = most of the time, 5 = all the time). Each subscale is summed to provide a total social anxiety score, ranging between 18-90. Higher values equate to more social anxiety. Reliability across all time points was good (18 items, α = .945).

#### Depression

The Beck Depression Inventory (BDI-II, ^48^) is a 21 item validated self-report instrument using a four-point scale ranging from 0 (symptom not present) to 3 (symptom very intense). Values are summed, greater values indicate more depression symptoms with a range from 0-63. Reliability across assessments was good (21 items, α = .825).

#### Quality of sleep

The Pittsburgh Sleep Quality Index (PSQI, ^52^) is a validated tool used for assessing sleep quality and disturbances over a one-month time interval. It consists of 19 self-rated questions that generate seven component scores: subjective sleep quality, sleep latency, sleep duration, habitual sleep efficiency, sleep disturbances, use of sleeping medication, and daytime dysfunction. These component scores are then summed to produce a global score ranging from 0 to 21, with higher scores indicating poorer sleep quality. Reliability assessed on 14 items across all time points was good, α = .71.

#### Physical activity

The International Physical Activity Questionnaire - Short Form (IPAQ-SF, ^54^) is a validated tool used to assess physical activity levels over the past seven days. It gathers information on activities in four domains: work, transportation, household chores, and leisure-time activities. Respondents report the frequency and duration of their physical activities, categorized by intensity (vigorous, moderate, and walking). The IPAQ-SF provides a summary of total physical activity in MET-minutes, a measure of energy requirements for activity undertaken, that are categorised as low, moderate, or high activity engagement.

#### Premenstrual screening

The Premenstrual Symptoms Screening Tool revised for adolescents (PSST-A, ^53^) is a validated questionnaire designed to assess premenstrual symptoms in young females. It includes questions that evaluate both physical and emotional symptoms experienced during the premenstrual phase of the menstrual cycle. Symptoms are scored on a scale from 0 to 4, with 0 indicating “not at all” and 4 indicating “extremely.” Reliability assessed on 14 items across assessments was good, α = .842. Items are recoded and summed according to symptom categories to provide three classifications of premenstrual symptoms severity: none/mild, moderate (indicative of premenstrual syndrome, PMS) and severe (indicative of premenstrual dysphoric disorder, PMDD).

#### Nutritional intake

was measured using a food diary over 4 days. Participants were asked to record everything they ate and drank, including brands, portion sizes and time of consumption. These data were entered into a nutritional analysis database ^55^ that estimated daily energy intake, and macro- and micro-nutrient intakes averaged over 4 days. The macronutrients were converted to a percentage of average energy intake using 4 calories per gram for carbohydrates and protein, 9 calories per gram for fat, and 8 calories per gram for alcohol.

### Behavioural tasks

#### Attentional Dot-Probe task

This measure is validated for measuring attentional bias in emotional disorders behaviourally ^49^, using emotionally valanced words (positive, negative and neutral) in both masked and unmasked conditions to alter awareness of stimulus emotional valence. Participants were positioned approximately 1 meter from a 19” LED monitor (native resolution 1280 x 1024 SXGA, brightness 250 cd/m2) on which white stimuli were presented (size 36 Aerial font) on a black background. PsychoPy ^56^ was used to present experimental stimuli, and responses collected by keyboard with 1000Hz polling rate. At the beginning of each trial, a fixation cross appeared on screen, followed with a word pair that was one of 30 positive-neutral, 30 negative-neutral and 30 neutral-neutral pairs, with words positioned at the top and bottom of the screen. Emotional word position was equally split across top and bottom. In the masked condition (90 trials), word pairs were presented for 17 ms, followed by a length and position matched mask of a nonsense letter string for 483 ms. In the unmasked condition (90 trials), word pairs were presented for 500 ms. This was followed by a probe of either one or Placebo stars that was congruent or incongruent with the emotional word position. The participant responded by pressing either ‘1’ or ‘2’ corresponding to the number of stars in the probe, terminating the trial. Frequency of number of stars and of position was equally split across all trials. A total of 180 trials were presented and randomised across masked and unmasked conditions, and valence stimulus pairs. Attentional vigilance was calculated from the response rate of correct responses by subtracting congruent response from incongruent responses. This was done for positive and negative stimuli in masked and unmasked conditions separately.

#### Reaction times

The Deary-Liewald Reaction Time task ^50^ is a short and simple stimulus-response task assessing reaction time in two tasks, Simple Reaction Time (SRT) and Four-Choice Reaction Time (CRT) tasks. Presented under the same conditions as described for the attentional dot probe task, participants press a button each time an ‘x’ is presented on screen. Mean and standard deviation measurements were taken for each participant on both the SRT and CRT tasks. In the SRT task, participants were required to press a key in response to a single stimulus. Conversely, the CRT task involved four stimuli, and participants had to press the key corresponding to the correct response. There were eight practice trials and twenty test trials for the SRT and eight practice trials and forty test trials for the CRT. This is an objective assessment of arousal and can indicate the emotional state of the participant ^50^.

### Gut microbiome

#### Stool sampling

Sampling kits were provided by BaseClear, the Netherlands including hygienic tools, stool catcher, collection tubes containing DNA/RNA Shield (Zymo Research, CA, USA) and secure return packaging with instructions. Samples were collected at home and kept at ambient room temperature until returned to the laboratory (within 1 month) for storage at − 80 °C.

#### Next generation sequencing: shallow shotgun metagenomics

All samples were shipped to Novogene, Cambridge, UK, for shallow shotgun metagenomic analysis^57^. This process involved several stages, including DNA extraction, quality control (QC), metagenomic library preparation, sequencing, and subsequent taxonomic and functional annotation. Samples that met QC thresholds, which included absence of contamination and sufficient volume, proceeded to library construction.

The library construction process involved randomly fragmenting genomic DNA into approximately 350 bp segments using a Covaris ultrasonic disruptor. These fragments were then end-repaired, A-tailed, and ligated with sequencing adapters. After this, the library was size-selected, PCR-amplified, purified, and quantified using Qubit and qPCR methods. Libraries that passed QC, including those with effective library concentrations exceeding 3 nM, were pooled according to target data output requirements, and subjected to shallow shotgun sequencing. Sequencing was performed on the NovaSeq X Plus Series platform, using paired-end 150 bp reads (PE150), and generated 6 GB of raw data per sample.

After sequencing, quality control was conducted using Fastp ^58^ to remove paired reads that contained adapter contamination, had more than 10% uncertain nucleotides, or contained more than 50% low-quality nucleotides (base quality <5). To further ensure data quality, clean reads were aligned to the host genome using Bowtie2 (Langmead & Salzberg, 2012) with the parameters --end-to-end, --sensitive, -I 200, -X 400, following previously established protocols ^65,70,71^.

The filtered clean data was assembled into contigs using MEGAHIT with meta-large presets ^67^. During the assembly process, scaftigs, which are scaffolds without ’N’ regions, were produced by breaking scaffolds at the ‘N’ junctions ^72^. Gene prediction was carried out using MetaGeneMark on scaffolds greater than or equal to 500 bp in length ^73^, and sequences shorter than 100 nucleotides were filtered out ^72^. To remove redundancy, the predicted genes were dereplicated using CD-HIT, creating a non-redundant gene catalogue^68^.

For taxonomic annotation, the predicted genes were aligned to the MicroNR database (https://www.ncbi.nlm.nih.gov), which contains sequences from bacteria, fungi, archaea, and viruses. The alignment was performed using DIAMOND ^60^, employing the blastp algorithm with parameters set to -e 1e-5 ^65^. The taxonomy of the sequences was assigned using the lowest common ancestor (LCA) algorithm, implemented in MEGAN ^63^. Species abundance was calculated by summing the gene abundances corresponding to each species ^62,70,74^.

Of the 178 samples returned by participants, 167 passed all QC thresholds and were available for analysis, with number of samples available by group at each time point and for each population specified in **Table 1**.

**Table 1.**
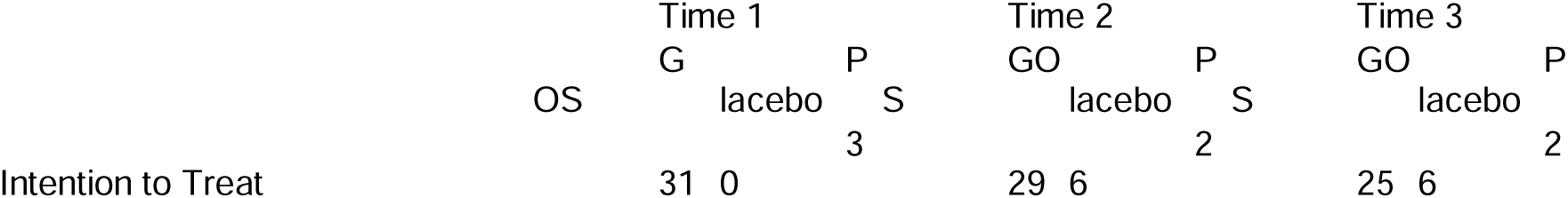
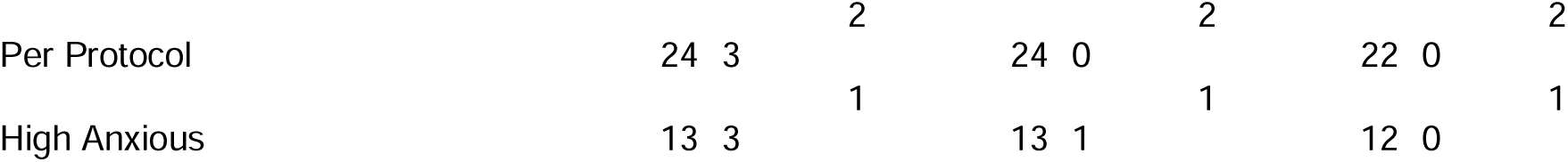
Number of available stool samples for metagenomic gut microbiome analysis by group, time, and population.

|  | Time 1 |  |  |  | Time 2 |  |  |  | Time 3 |  |
| --- | --- | --- | --- | --- | --- | --- | --- | --- | --- | --- |
|  | OS | G | P | S | GO | P | S | GO | P | S |
|  |  | lacebo |  |  | lacebo |  |  | lacebo |  |  |
| Intention to Treat |  | 31 | 0 | 3 | 29 | 6 | 2 | 25 | 6 | 2 |

|  |  |  |  |  |  |  |
| --- | --- | --- | --- | --- | --- | --- |
|  |  | 2 |  | 2 |  | 2 |
| Per Protocol | 24 3 |  | 24 0 |  | 22 0 |  |
|  |  | 1 |  | 1 |  | 1 |
| High Anxious | 13 3 |  | 13 1 |  | 12 0 |  |

#### Bioinformatic analysis of sequenced stool samples

Bioinformatics analysis of the DNA sequenced from stool samples aimed to determine if the supplement intervention had effects on gut microbiome composition with predictions that GOS would increase *Bifidobacterium* abundance in comparison to placebo. To investigate the gut microbiome composition diversity metrics were calculated on all available annotated metagenomic taxa (n = 17130) for each sample, including Abundance-based Coverage Estimator (ACE) and Chao1 to estimate richness, Shannon and Simpson metrics to estimate richness and evenness, counts of observed species, and Goods coverage to estimate community diversity.

Supplement intervention effects on the gut microbiome were then investigated with a two-step differential abundance analysis. First the R package Microbiome Multivariable Association with Linear Models ^75^, (MaAsLin2, version 1.15.1) was used to determine if there was an association with supplement exposure (e.g., GOS versus placebo) on gut microbiota at the genus level over time on the gut microbiome. Absolute abundances of metagenomic taxa were filtered at a 100% prevalence across all samples to retain n = 195 core taxa.

Analysis was normalised using a Centred Log-Ratio (CLR) transformation to address compositionality. Fixed effects were specified for supplement group (GOS versus placebo) and time (baseline, endline, follow up), and an interaction term of supplement group*time.

Second, the R package Analysis of Compositions of Microbiomes with Bias Correction 2 ^76^, (ANCOM-BC2, version 2.4.0), selected for stringent bias correction analysis, was used to quantify the magnitude of change in microbiota at the genus level over time in each supplement group separately. Core taxa were identified across all samples using prevalence of 97%, n = 591. To address the presence of zero counts in compositional data, pseudo-count sensitivity analysis was enabled. Additionally, taxa exhibiting consistent zeros across samples were flagged as structural zeros, and a negative lower bound adjustment applied for these cases. Mixed directional False Discovery Rate (mdFDR) was employed to account for multiple comparisons across taxa and pairwise group tests.

### MRI scanning

#### Anatomical acquisition

MRI data were acquired on a Siemens 3T Magneton TIM Trio scanner with a 32-channel head coil. Sagittal T1-weighted Magnetization Prepared Rapid Acquisition Gradient Echo (MPRAGE) images were acquired with parameters TR = 1900 ms, TE = 3030 and dwell time 1500 ms, 1 mm slices in field of view (FOV) resolution 256 x 256 over 5 minutes acquisition time. T2*-weighted Turbo Spin Echo (TSE) images were acquired in the coronal and axial planes with TR = 3500 ms, TE = 93 ms, dwell time 7100 ms, GRAPPA acceleration factor = 2, and FOV of 179 x 256 in 4 mm slices. Structural images were reconstructed to guide voxel placement for ^1^H-MRS acquisition.

#### ^1^H-MRS data acquisition

^1^H-MRS in vivo spectra were acquired in three volumes of interest (VOIs), left dorsolateral prefrontal cortex (DLPFC), anterior cingulate cortex (ACC) and left inferior occipital gyrus (IOG). A 2 cm³ voxel was manually centred within each VOI, guided by sagittal T1-weighted images.

Spectra were acquired using the Spin Echo full Intensity-Acquired Localized spectroscopy sequence, SPECIAL ^77^. Water suppressed spectra were acquired with TR = 3200 ms, TE 8.5 ms, flip angle of 90° over 192 averages. Water unsuppressed spectra were acquired beforehand over 16 averages in the same region. To minimise unwanted signals outside the voxel boundaries, saturation bands were manually aligned along the six outer planes of each voxel before acquisition. **Figure 3** displays a representation of voxel placement over each region of interest.

**Figure 3.**
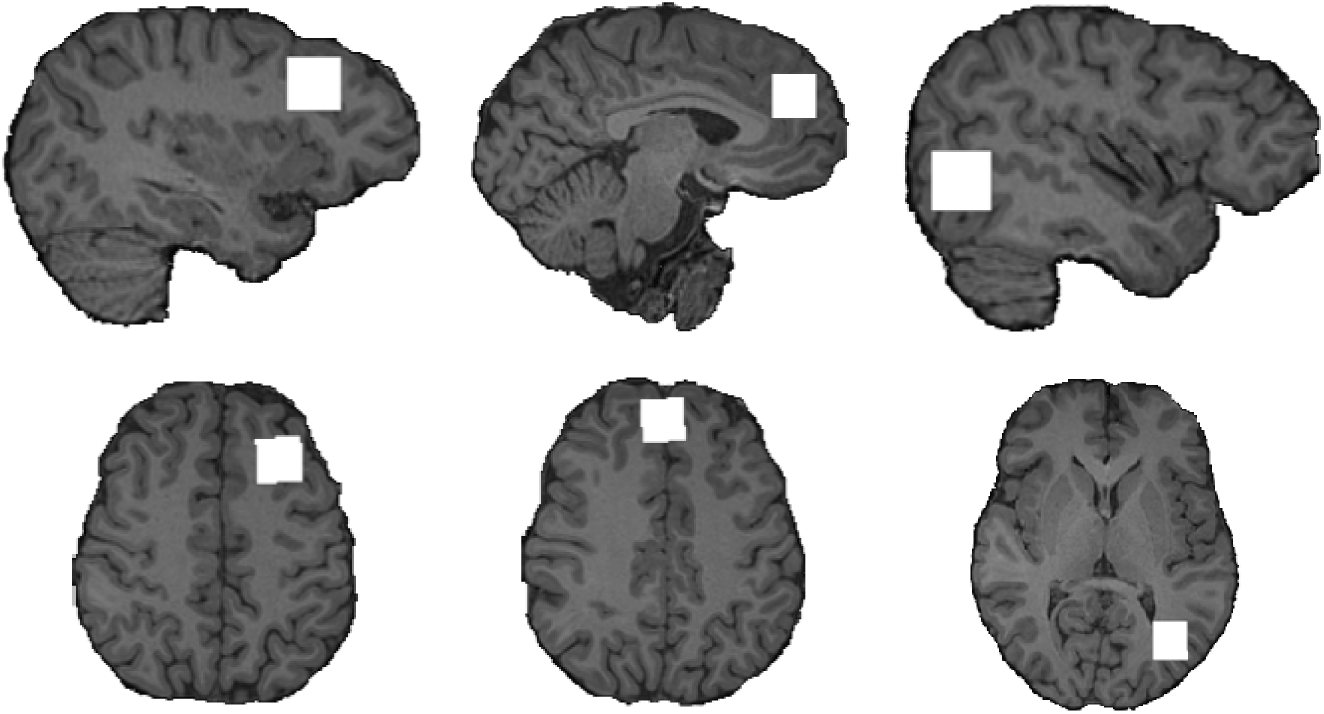
A representation of voxel locations over each region, shown as white squares, all 2 cm^3^. The most lateral sagittal edge and superior axial edge are shown (from left to right) for the dorsolateral prefrontal cortex VOI, anterior cingulate gyrus VOI, and inferior occipital gyrus VOI. Voxel locations were based on a reference image and agreed upon by two experienced MRI operators.

#### ^1^H-MRS data pre-processing

For each VOI, processing functions in the FID-A toolbox ^78^ in MATLAB (The Mathworks, Natick, MA) were used to remove motion corrupted averages and correct frequency drift. Voxel tissue was apportioned for cerebrospinal fluid (CSF) correction^79^ using the segment function of the Gannet toolbox ^80^ which uses the Statistical Parametric Mapping toolbox (SPM12, MATLAB, The Mathworks, Natick, MA) for co-registration and segmentation ^81^. Detailed formulas for tissue correction are available at <u>Gannet documentation</u>.

Cleaned averaged spectra were analysed in LCModel ^82^ to estimate relative concentrations of GABA and glutamate in each VOI, referenced to the water peak of the VOI. Spectral quality was assessed through: (i) line width of the water peak (full width at half maximum), excluding widths >13 Hz; (ii) signal-to-noise ratio (SNR), defined as the NAA peak height (1.8–2.2 ppm) divided by noise (averaged signal from -2 to 0 ppm), with SNR <150 excluded; (iii) visual confirmation of NAA, Cr, Cho, and glutamate peaks at expected ppm values; and (iv) Cramer-Rao lower bounds (CRLBs), with thresholds of >29 for GABA and >15 for glutamate. These stringent criteria ensured consistency across timepoints and VOIs.

### Statistical analysis

The statistical analysis plan was written for intention to treat (ITT), per protocol (PP) and high anxious subgroup and performed in R ^83^. Missing data was not imputed.

Ancova models were used to test supplement intervention effects by comparing daily GOS exposure versus daily placebo exposure for 28 days in separate groups on primary (trait anxiety), secondary (brain measures, gut microbiome composition and diversity measures) and tertiary outcomes (other psychological, behavioural and nutrition measures) separately. Baseline measures of the outcomes were included as covariates in addition to quality of sleep, physical activity, menstrual cycle phase and BMI. For nutritional analysis, diet type and energy intake in calories were used as covariates.

Linear mixed models were used to explore longevity of effects over the study period estimating fixed effects of the supplement group (GOS or placebo) and time (supplement intervention period, contrast T1 – T2; and follow up, contrast T1-T3) on primary and secondary outcomes. Random effects were specified at the participant level, and covariates the same as for ancova. Simple effect contrasts were reported over time within groups and between groups at time 1, 2 and 3 to illustrate pattens of effects. Models will be considered significant at *p* < .05 and considered trend level around *p* < .1.

Differential abundance analysis of the gut microbiome was performed, following a specialised analysis plan to accommodate the volume and compositional nature of the data to determine which gut microbiota differ over time due to GOS supplementation. MaAsLin2 was used to assess the effects of GOS versus placebo groups across all time points. The resulting coefficients represented the size and direction of the effect of the supplement intervention on the relative abundance of each taxon, accounting for compositionality. P-values were used to assess the statistical significance of these associations, while q-values were calculated to account for adjustments due to multiple testing and control for false discovery.

To further examine within-group changes over time, ANCOMBC2 was applied separately to the GOS and placebo groups. This method identified which microbiota was differentially abundant over time, using the W-statistic to reflect the strength of the association, p-values for significance and q-values for mdFDR. Differential abundance was quantified using log fold change (LFC), which represents the logarithmic ratio of relative abundance changes between time points, providing a measure of the direction and scale of change in microbial taxa.

## Results

### Descriptive characteristics and compliance

An overview of demographic variables for all enrolled participants in presented in **Table 2**. 61 participants provided endline measures for the primary outcome (trait anxiety). The average supplement intervention period was 28.1 days (*SD* = 4.3, range 17-43 days; group GOS *M* = 28.6 days, *SD* = 4.92; placebo group *M* = 27.5 days, *SD* = 3.53). In the intervention period, an average of 25.3 supplements were consumed (*SD* = 4.29, range 5-28; group GOS *M* = 24.9 days, *SD* = 5; placebo group *M* = 25.8 days, *SD* = 3.43). A compliance score was calculated by dividing the number of supplements consumed by days in the intervention period. Those with a compliance score of 80 and above were included in the per-protocol analysis if they also provided the primary measure at baseline and follow up. This produced a sample size of n = 52, n = 26 in each group at endline. Sensitivity analysis was performed on a high anxious subgroup. This included participants scoring higher than the sample median on the trait subscale of the STAI at baseline, giving a total sample of n = 26, n = 13 per group at endline. Participants were asked which supplement they believed they had consumed at endline and follow up, with more participants reporting placebo at both endline ( X^2^ (1, *N* = 55) = 11.36, *p* < .001) and follow up ( χ^2^ (1, *N* = 51) = 10.37, *p* = .001).

**Table 2.** Overview of demographic and covariate variables for all participants (n = 83, ITT population).

| Time point | 1 |  | 2 |  | 3 |  |
| --- | --- | --- | --- | --- | --- | --- |
| Demographics | GOS, N = 42<sup>1</sup> | Placebo, N = 41<sup>1</sup> | GOS, N = 32<sup>1</sup> | Placebo, N = 30<sup>1</sup> | GOS, N = 29<sup>1</sup> | Placebo, N = 29<sup>1</sup> |
| Age | 20.08<br>(2.63) [2] | 20.56<br>(2.61) [4] | 20.45<br>(2.70) | 20.79<br>(2.57) | 20.57<br>(2.76) | 20.89<br>(2.55) |
| Trait anxiety | 47 (11) | 43 (9) [1] | 45 (10) [1] | 42 (10) | 43 (10) | 42 (10) |
| Quality of Sleep | 7.17<br>(3.12) | 6.63 (3.03) | 6.5 (3.9) | 5.1 (2.3) | 6.14<br>(3.24) | 5.07 (2.62) |
| Physical activity |  |  |  |  |  |  |
| High | 2 (4.8%) | 3 (7.3%) | 1 (3.1%) | 3 (10%) | 1 (3.4%) | 1 (3.4%) |
| Moderate | 37 (88%) | 32 (78%) | 26 (81%) | 22 (73%) | 24 (83%) | 18 (62%) |
| Low | 3 (7.1%) | 6 (15%) | 5 (16%) | 5 (17%) | 4 (14%) | 10 (34%) |
| Menstrual phase |  |  |  |  |  |  |
| Follicular | 16 (47%) | 13 (37%) | 10 (40%) | 13 (48%) | 10 (45%) | 14 (58%) |
| Luteal | 18 (53%)<br>[8] | 22 (63%)<br>[6] | 15 (60%)<br>[7] | 14 (52%)<br>[3] | 12 (55%)<br>[7] | 10 (42%)<br>[5] |
| Diet |  |  |  |  |  |  |
| Omnivore | 39 (93%) | 38 (93%) | 30 (94%) | 28 (93%) | 27 (93%) | 27 (93%) |
| Vegetarian | 3 (7.1%) | 3 (7.3%) | 2 (6.3%) | 2 (6.7%) | 2 (6.9%) | 2 (6.9%) |
| BMI | 23.8 (4.1) | 22.4 (3.5)<br>[1] | 23.7 (4.2) | 22.3 (3.5) | 23.8 (4.3) | 22.4 (3.5) |
| Compliance | 89 (18)<br>[11] | 94 (10)<br>[12] | 89 (18) [1] | 94 (10) [1] | 90 (18) | 94 (10) |
| Supplements consumed | 24.9 (5.0)<br>[11] | 25.8 (3.4)<br>[11] | 24.9 (5.0) | 25.8 (3.4) | 24.9 (5.2) | 25.7 (3.5) |
| Guess |  |  |  |  |  |  |
| Placebo |  |  | 22 (79%) | 18 (67%) | 20 (80%) | 17 (65%) |
| GOS |  |  | 6 (21%)<br>[4] | 9 (33%)<br>[3] | 5 (20%)<br>[4] | 9 (35%)<br>[3] |

<sup>1</sup>Mean (SD); n (%); [missing]

### Primary outcome: Trait anxiety

On the primary outcome of trait anxiety, there were no baseline group differences, and the distribution of trait anxiety scores was normal. The covariate quality of sleep was related to anxiety outcomes, with better quality of sleep associated with lower anxiety (β = 4.69, *SE* = 2.42, *p* < 0.001. Accounting for this, ancova models of supplement intervention effects found no significant differences between GOS and Placebo groups at endline in ITT (*F*(3,57) = 0.59, *p* = .443), PP (*F*(3,48) = 0.62, *p* = .433) or high anxious populations (*F*(2,23) = 0.19, *p* = .665). Mixed model analysis of ITT population found that poorer quality of sleep was associated with higher anxiety levels (β = 3.06, *SE* = 0.69, *p* < .001). Accounting for this, there was a trend level supplement group difference in the follow up period (T3-T1, β = 2.89, *SE* = 1.57, *p* = .069). Simple effects analysis of both between group effects (comparing supplement groups at each time point), and within group effects (comparing responses at each time point for each group) showed that GOS group reduced anxiety levels over time, significantly at follow up (β = -2.68, *SE* = 1.12, *p* = .034), which was not evident in placebo group (β = 0.21, *SE* = 1.15, *p* = .96), **Figure 4**.

**Figure 4.**
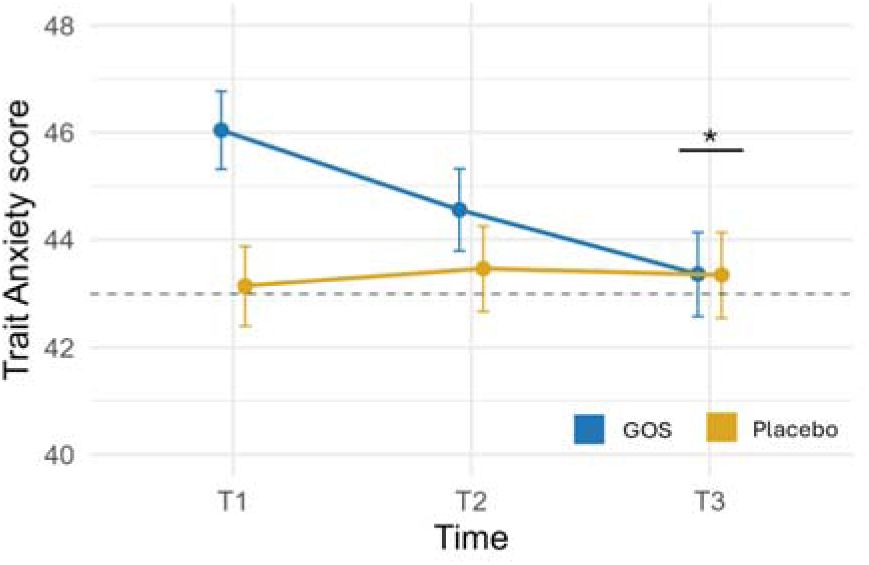
Results of trait anxiety longevity analysis by mixed models in ITT population. Trait anxiety reduces over time in the GOS group and is significant at T3 Dashed horizontal line shows boundary for inclusion in high anxious subgroup. Point estimates are estimated marginal means (EMM) from the mixed model analysis and error bars are standard error of EMM. \**p* >0.05.

### Secondary outcome: Neurochemical effects

The neurochemicals under investigation, GABA and glutamate were obtained from 3 regions of the brain, DLPFC, ACC and IOG. There were no baseline group differences, nor influential covariates on GABA levels. GABA distribution was non-normal thus a gamma distribution was fit to model effects in a generalised linear model. In the ITT population, analysis of intervention effects on GABA found no supplement group difference but a regional difference was evident, *F*(2,141) = 5.61, *p* = .005. Examining the group effects within each region found a trend level group difference in the IOG where GABA levels were lower in GOS group versus placebo (β = -0.079, *SE* = 0.04, *p* = .053, GOS group *EMM* = 1.50, *SE* = 0.65; placebo group *EMM* = 1.70. *SE* = 0.81), **Figure 5**, panel A. A similar trend level pattern was found in the PP population, but not the high anxious population, **Figure 5**, panel B. In the high anxious population, simple effects analysis of the regions found a trend to reduced GABA levels in the DLPFC in GOS group versus placebo (β = -0.125, *SE* = 0.02, *p* = .088, GOS group *EMM* = 1.29, *SE* = 0.09; placebo group *EMM* = 1.54. *SE* = 0.12), **Figure 5**, panel C.

**Figure 5.**
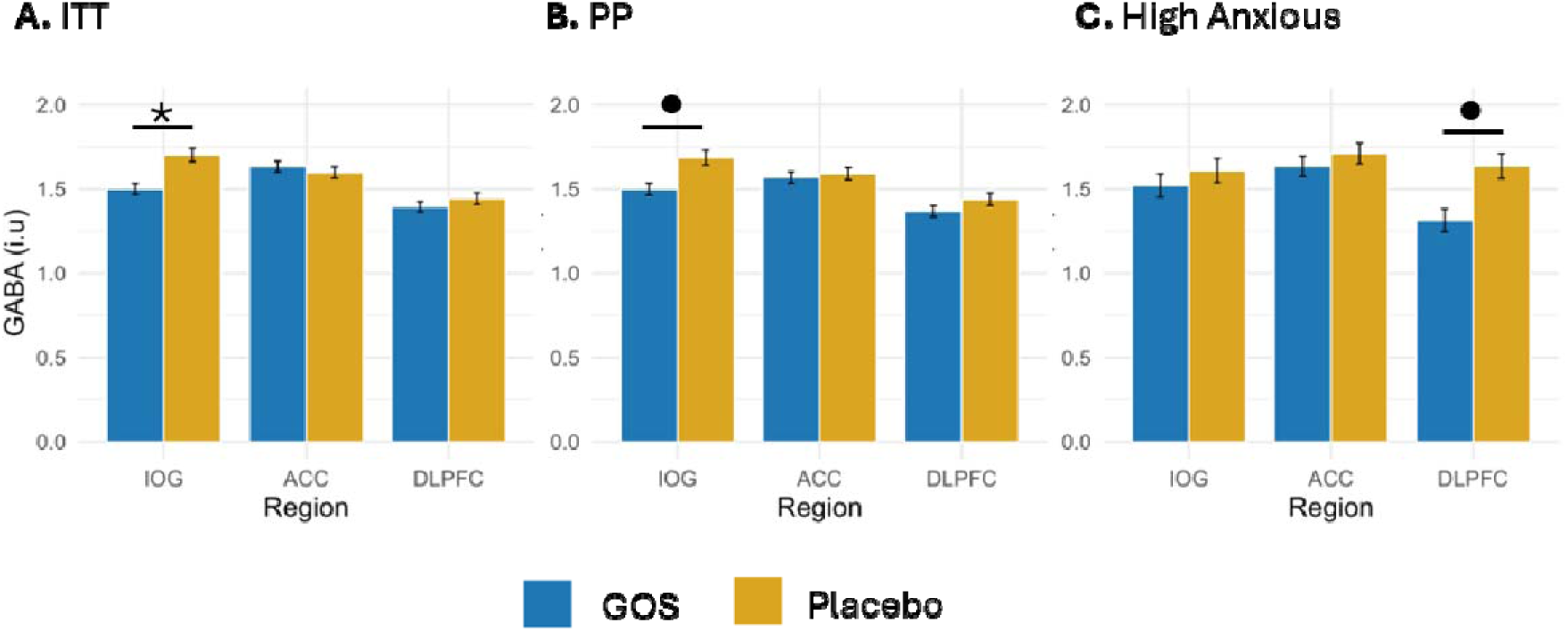
Results of GABA outcomes following supplement intervention. In panel A, GABA levels are lower in the IOG in GOS group compared to placebo group for the ITT population. The same effect is seen in panel B for the PP population. In panel C, GABA levels are lower in the DLPFC in the GOS group compared to placebo group. At trend level. Bars plot estimated marginal means (EMM) from ANCOVA, and error bars are standard error of EMM. \**p* >0.05, ^●^ *p* > 0.1.

Due to the identified regional variations in GABA levels, mixed models including all three time points were run for each region separately. For ITT and PP populations in both the ACC and DLPFC there were no group differences in GABA levels. In the IOG there were effects following intervention (β = -0.153, *SE* = 0.04, *p* < .001) and at follow up longevity effects (β = -0.171, *SE* = 0.04, *p* < .001). Simple effects analysis showed this was due to lower GABA levels in GOS group compared to placebo group at endline T2 (β = -0.090, *SE* = 0.04, *p* = .046) and follow up (T3) (β = -0.109, *SE* = 0.04, *p* = .016); driven by reducing GABA levels in GOS group), **Figure 6**, panel A. In the high anxious population, the intervention effect identified by ANCOVA in the DLPFC was evident (β = -0.169, *SE* = 0.07, *p* = .016) but was not maintained at follow up (β = -0.110, *SE* = 0.07, *p* = .115), **Figure 6**, panel B. However, in the ACC, there was a group difference at follow up (T3) (β = -0.111, *SE* = 0.04, *p* = .002) where GABA levels were lower in GOS group *EMM* = 1.51, *SE* = 0.07 than placebo group *EMM* = 1.84. *SE* = 0.11, **Figure 6**, panel C.

**Figure 6.**
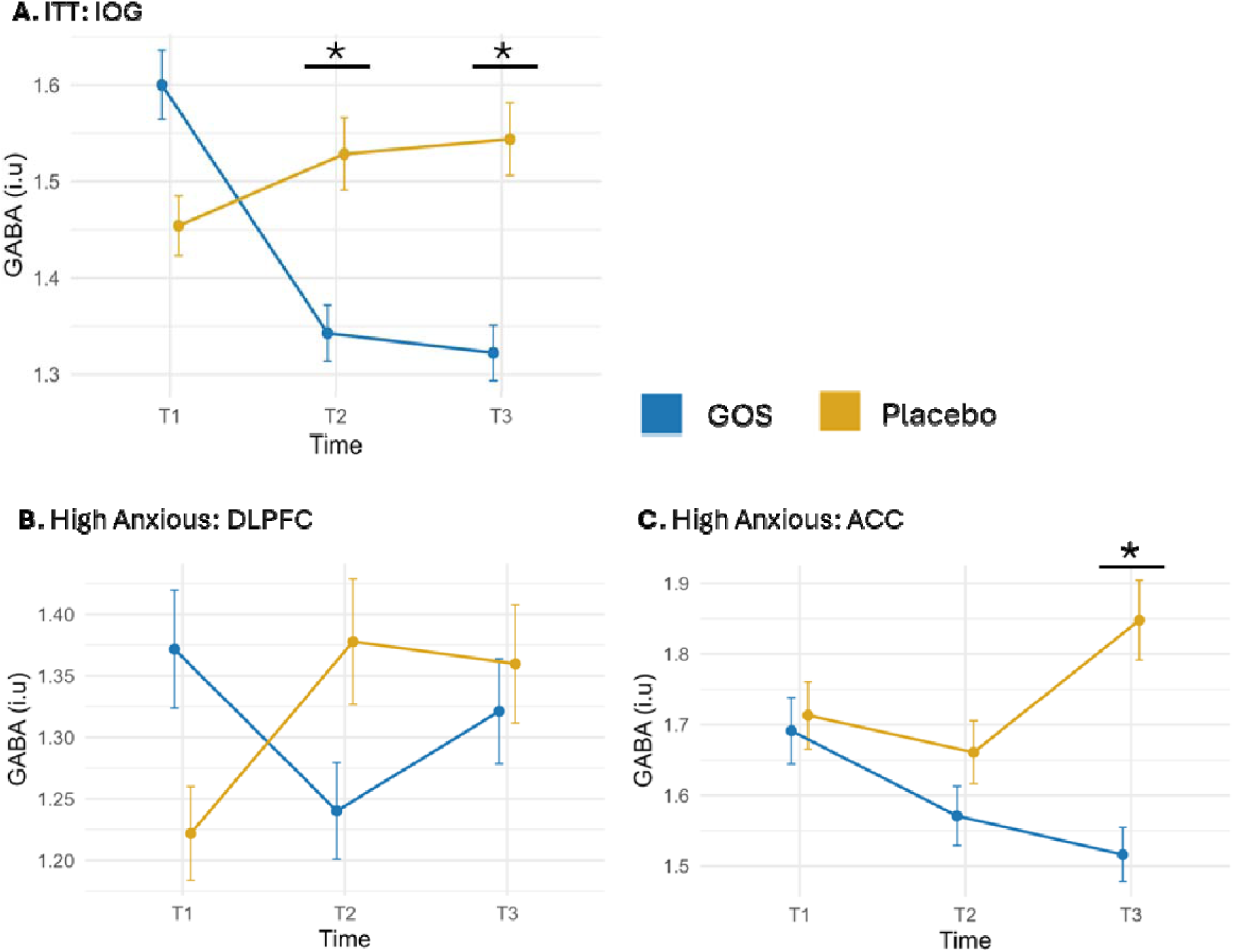
Results of GABA longevity analysis by mixed models. In panel A, the GABA response in the ITT population is shown. GABA levels reduce in the GOS group compared to placebo in the IOG at T2 following intervention is maintained at T3. Panels B and C show GABA response in the high anxious population. In panel B, GABA is lower at T2 in the GOS group compared to placebo, and in panel C, GABA is significantly lower at T3 in the GOS group in the ACC, compared to placebo. Point estimates are estimated marginal means (EMM) from mixed model analysis and error bars are standard error of the EMM. \**p* < 0.05.

Regarding glutamate, there were differing levels at baseline by region (ACC had higher levels compared to DLPFC and IOG which had similar levels) and there were no baseline group differences between GOS and placebo, nor influential covariates on glutamate levels. Glutamate distribution was non-normal thus a gamma distribution was fit to model effects in a generalised linear model. For ITT, PP and high anxious populations, there were no intervention effects on glutamate levels.

In the mixed model analysis, glutamate levels were lower at follow up (T3) in the DLPFC of the ITT population for GOS group compared to placebo group (β = -0.011, *SE* = 0.005, *p* = .050, GOS group *EMM* = 5.68, *SE* = 0.14; placebo group *EMM* = 6.09. *SE* = 0.15). For the PP population in the ACC there was trend level lower glutamate levels for GOS group compared to placebo group at follow up (β = 0.54, *SE* = 0.32, *p* = .090, group GOS *EMM* = 7.78, *SE* = 0.17; placebo group *EMM* = 8.08. *SE* = 0.17), and a similar pattern in the IOG (β = 0.40, *SE* = 0.25, *p* = .10, group GOS *EMM* = 4.93, *SE* = 0.13; placebo group *EMM* = 5.06. *SE* = 0.13). Reduced glutamate levels for group GOS at follow up were also observed in the high anxious population in the ACC (β = 0.92, *SE* = 0.40, *p* = .02, group GOS *EMM* = 7.30, *SE* = 0.23; placebo group *EMM* = 8.26. *SE* = 0.26), but not in the DLPFC or IOG.

The balance of glutamate and GABA, or excitation/inhibition ratio (E/I ratio) is an important index in determining if there are alterations in cortical responsiveness across the regions. At baseline, E/I ratio differed by region and was lower in the IOG compared to both the ACC and DLPFC which has similar levels. There were no baseline group differences in E/I ratio, nor influential covariates. E/I ratio distribution was normal thus a gaussian distribution was fit to model effects in a generalised linear model. No intervention effects were found in the ITT or PP populations. In the high anxious population, simple effects analysis by region showed increased E/I ratios in the DLPFC (β = 0.69, *SE* = 0.32, *p* = .040, group GOS *EMM* = 4.73, *SE* = 0.21; placebo group *EMM* = 4.04. *SE* = 0.20), reflecting the decrease in GABA. The mixed model analysis found trend level intervention and follow up effects for the ITT population in the ACC (β = -0.014, *SE* = 0.01, *p* = .079 and β = -0.014, *SE* = 0.01, *p* = .094, respectively) driven by an increase in E/I ratio in placebo group. A similar pattern was seen in the PP population but not the high anxious population.

### Secondary outcome: Gut microbiome

On the diversity of the gut microbiome, the metrics calculated from all sequenced taxa (n = 17130) included ACE, Chao1, Shannon, Simpson, observed counts and Goods coverage. Across all populations, there were no group differences at baseline for any diversity measure. Similarly, there were no differences between GOS and placebo identified by ANCOVA in the ITT, PP, or high anxious populations, or in the mixed model analysis of longevity effects.

Differential effects of GOS and placebo were on gut microbiome composition at the genus level were contrasted in a mixed MaAsLin model with fixed effects specified as the supplement intervention group, time, and an interaction term of supplement intervention group and time to contrast GOS versus placebo at each time point. We predicted that GOS would increase *Bifidobacterium* abundance. In the ITT population the model found a trend level main effect of supplement intervention group, β = -0.46, *SE* = 0.27, *p* = .099; a main effect of time, β = 0.51, *SE* = 0.25, *p* = .048, and interaction, β = -0.79, *SE* = 0.39, *p* = .042. Note, mdFDR corrections reported q-values all greater than .739, giving a high probability of Type I error within the sample. However, given our predictions for GOS supplementation to increase the abundance of *Bifidobacterium,* we proceeded with further analyses, noting that other significant changes are less likely to be true effects as restricted by the available sample size. In the PP population the model found similar results, a trend level main effect of supplement group, β = -0.51, *SE* = 0.30, *p* = .098; a main effect of time, β = 0.62, *SE* = 0.28, *p* = .030, and interaction, β = -0.88, *SE* = 0.42, *p* = .041. However, this pattern was not found in the high anxious population; main effect of supplement group, β = -0.19, *SE* = 0.46, *p* = .671; a main effect of time, β = 0.67, *SE* = 0.47, *p* = .112, and interaction, β = -0.56, *SE* = 0.64, *p* = .385.

The magnitude of change in taxa over time was established with ANCOMBC2 models of genus level in each supplement group separately. Here we report (i) which taxa showed a significant change over all time points (baseline, endline an follow up) defined by the W statistic > 1 and *p* < 0.05, and (ii) determining the direction and influential time points with LFC of these taxa. Full LFC analysis is available in supplementary **Table S1**.

#### Differential abundance in GOS group

In the ITT population of GOS group at the genus level, differentially abundant taxa included *Bifidobacterium* (W = 8.05, *p* = .001, *q* = 0.321), *Paraclostridium* (W = 4.62, *p* = .025, *q* = 0.581), *Rhizopus* (W = 4.68, *p* = .024, *q* = 0.581), and *Gardnerella* (W = 6.92, *p* = .003, *q* = 0.321). **Figure 7**, panel A shows *Bifidobacterium* increasing at time two by 87.39%, following intervention, and decreasing at time 3 by 31.75%, at follow up a pattern consistent and specific to the period in which GOS was consumed, while the other taxa show increases either at both time 2 and time 3, or decreases (Table S1).

**Figure 7.**
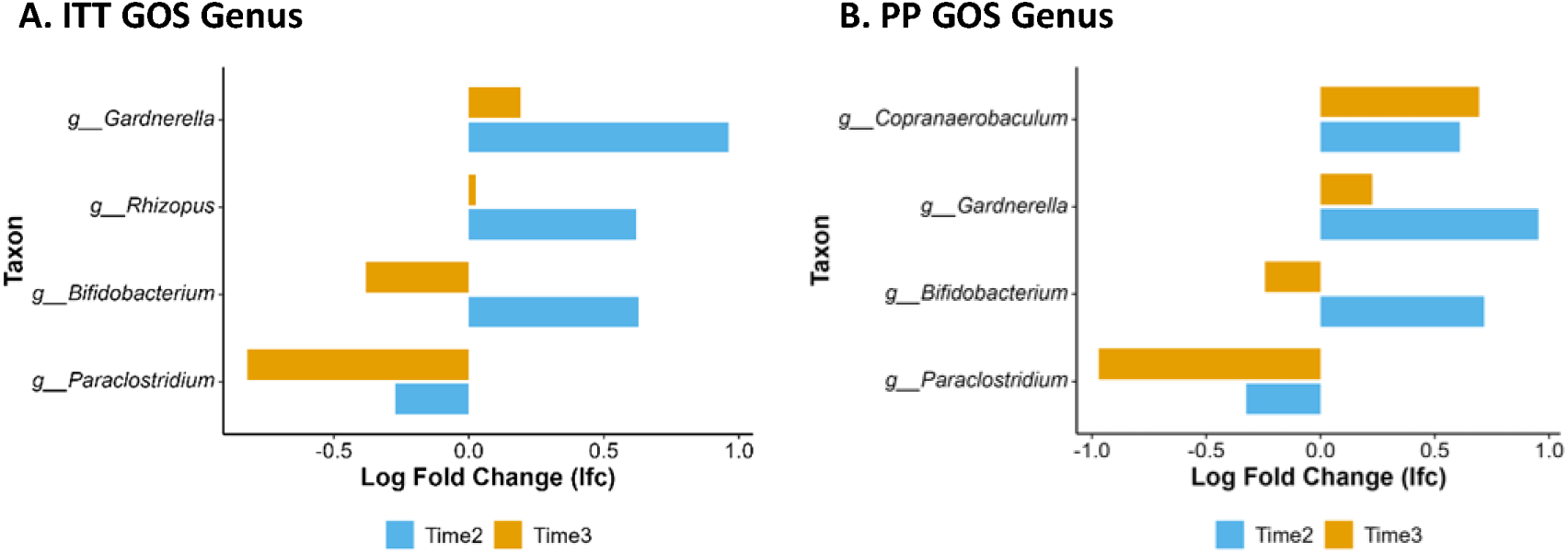
Log fold change at time 2 and time 3 for each differentially abundant microbe identified in the GOS group at genus level of ITT and PP populations.

In the PP population of the GOS group at the genus level, differentially abundant taxa included *Bifidobacterium* (W = 6.22, *p =* 0.007, *q =* 0.879), *Paraclostridium* (W = 4.90, *p =* 0.021, *q =* 0.965), *Gardnerella* (W = 5.85, *p =* 0.009, *q =* 0.879), and *Copranaerobaculum* (W = 4.18, *p =* 0.039, *q =* 0.965). **Figure 7**, panel B shows an increase in *Bifidobacterium* at time 2 by 104.83% and decrease at time 3 by 21.57% consistent with the period of GOS supplementation. While the others either increase or decrease at both time 2 and time 3.

#### Differential abundance in placebo group

In the ITT population of the Placebo group at the genus level, there were no significant changes in taxa. In the absence of other differentially abundant taxa, results here show stability in the gut microbiome of the placebo group over time, and no specific growth or reduction of taxa due to the maltodextrin placebo. It also illustrates no significant transient or environmental exposures during the study period, proving confidence in the success of the supplement intervention protocol and illustration of differential abundance effects in the GOS group. An illustrative overview of the differential effects of GOS versus placebo on the scaled and normalised mean absolute abundance of *Bifidobacterium* is shown in **Figure 8**.

**Figure 8.**
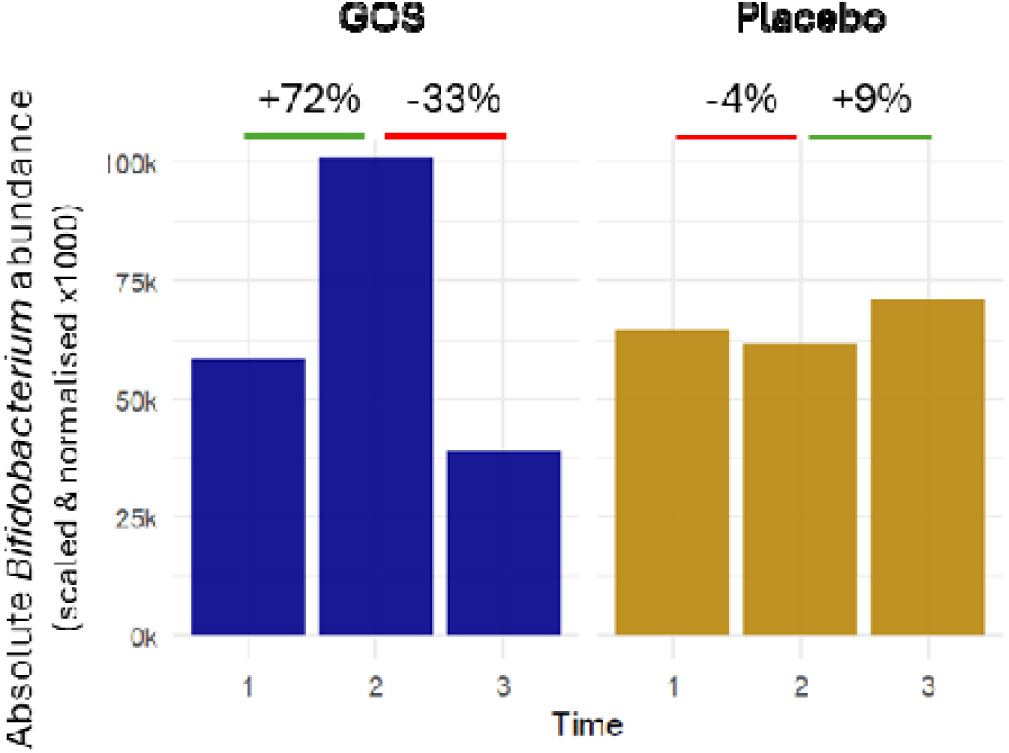
Scaled and normalised mean absolute abundance of *Bifidobacterium* over time in the GOS and placebo groups in the PP population. Percentage change is calculated from mean absolute abundance in reference to Time 1 in each group separately.

### Tertiary outcomes

For the psychological variables social anxiety and depression there were no baseline differences between groups and poorer quality of sleep was associated with both higher social anxiety and depression scores, yet there were no changes following supplement intervention in any population; Social anxiety, ITT *F*(3,57) = 0.13, *p* = .725; depression, ITT *F*(3,57) = 0.64, *p* = .426.

For the assessment of attentional bias to emotional stimuli in the dot-probe task, it is expected that emotional biases will become evident to positive and negative stimuli (in this case words) by scores greater than zero. The larger the value from zero, the greater the effect of the emotional stimuli (e.g., someone who held a negative disposition would spend more time drawn to a negative word, and the bias score would be larger). Where bias scores are less than zero, this indicates that the emotional stimuli have not been noticed. Mean emotional bias scores at baseline were close to zero for both groups and did not differ between groups. There was no change in scores following supplement intervention, *F*(8,227) = 0.01, *p* = .917.

In the behavioural simple reaction time task, mean reaction times were significantly quicker in GOS group compared to placebo group (*p* = 0.003) at baseline. Individual trial reaction times were transformed to the inverse to calculate reaction rate, and a gaussian distribution fit to the data. Higher physical activity levels were related to slower reaction rates and participation during the luteal menstrual phase was associated with quicker reactions compared to participation during the follicular phase. Accounting for this, there was no group difference at T2, following supplement intervention in the ITT or PP populations. In the high anxious population, there was a difference following supplement intervention where group GOS were quicker to respond than placebo group (*F*(1,448) = 4.41, *p* = .036; GOS group *EMM* = 0.183, *SE* = 0.001, placebo group *EMM* = 0.181, *SE* = 0.001, **Figure 9**, panel A).

**Figure 9.**
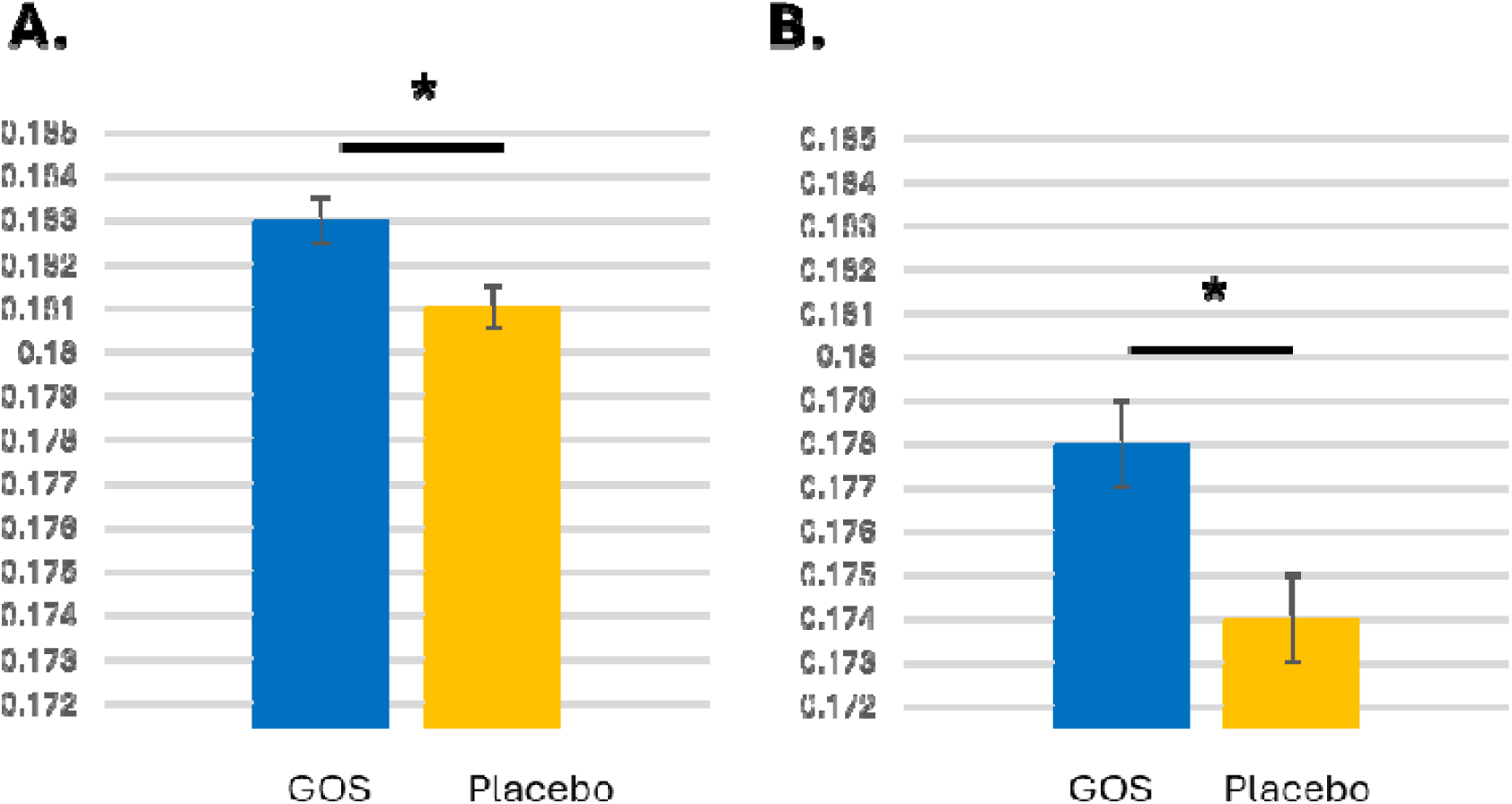
GOS versus Placebo intervention outcomes in high anxious participants on reaction rates in the simple reaction time task (panel A) and choice reaction time task (panel B). Higher values (y-axis, reaction rate, ms) indicate quicker reactions. In both tasks, simple and choice, those in the GOS group improved speed of response in contrast to those in the placebo group. *\* p* < 0.05.

In the behavioural choice reaction time task, mean reaction times were significantly quicker in GOS group compared to placebo group (p = 0.001) at baseline. The inverse transform was applied to individual trial reaction times to calculate reaction rates, and a gaussian distribution fit to the data. Higher physical activity levels were related to slower reaction rates and participation during the luteal menstrual phase was associated with quicker reactions compared to participation during the follicular phase. Poorer quality of sleep was also related to quicker reactions. Accounting for this, there was no group difference at T2 in the ITT population. However there were group differences in the PP and high anxious populations, with GOS group quicker than placebo group; PP: (*F*(1,1390) = 3.87, *p* = .049; group GOS *EMM* = 0.176, *SE* <0.001, placebo group *EMM* = 0.175, *SE* < 0.001), high anxious: (*F*(1,599) = 15.88, *p* < .001; group GOS *EMM* = 0.178, *SE* = 0.001, placebo group *EMM* = 0.174, *SE* = 0.001, **Figure 9** panel B). Results for supplement intervention effects in each population for each outcome can be found in Table S2.

### Tertiary outcome: Nutrition variables

Nutrient intake was measured using the average self-reported food and drink intake over 4 days at baseline, endline and follow up. This was quantified as average energy intake (in calories), and individual macronutrients (carbohydrates, protein, and fat) were quantified as a percentage of energy intake. Fiber and added sugars were also examined (quantified as grams). For the ITT population, nutrient intake data was available from 29 participants in the GOS group, and 31 participants in the placebo group.

In the mixed model analysis, for average energy intake, BMI, diet, and trait anxiety scores were examined as covariates. Higher trait anxiety scores were linked to lower energy intakes β = -111.31, *SE* = 44.47, *p* = .014. After accounting for this, no further effects of supplement group were observed at endline or follow-up. For the remaining nutrient intakes, BMI, diet, and energy intake were considered as covariates. In the case of carbohydrates, there were no covariate influences and no supplement intervention or follow-up effects. For protein higher energy intake was associated with lower protein intake, β = -0.059, *SE* = - 0.016, *p* < .001, and vegetarian diets also reduced protein intake β = -0.307, *SE* = 0.116, *p* = .010. However, there were no supplement intervention or follow-up effects. Regarding fat, higher energy intake was associated with higher fat intake, β = 1.18, *SE* = 0.53, *p* = .028, but again, there were no supplement intervention or follow-up effects. For fibre, higher energy intake was associated with increased fibre intake, β = 0.526, *SE* = 0.053, *p* < .001, though no supplement intervention or follow-up effects were found. Similarly, for free (added) sugars, higher energy intake correlated with higher free sugar intake, β = 0.821, *SE* = 0.157, *p* < .001, and after accounting for this, there were no supplement intervention or follow-up effects.

## Discussion

The present study investigated the effects of galacto-oligosaccharides (GOS) supplementation on trait anxiety, neurochemistry, gut microbiome composition, cognition, and nutritional outcomes in healthy young adult females. Although our primary hypothesis that GOS would reduce trait anxiety after supplement intervention, we observed no significant differences between the GOS and placebo groups. However, a trend-level reduction in anxiety was detected at follow-up, suggesting that psychological effects of GOS may require a longer duration to manifest.

Our secondary analyses revealed more pronounced effects in neurochemical and microbiome outcomes. While we predicted brain GABA levels would increase with GOS supplementation, we found the opposite to be true. Specifically, GOS supplementation resulted in a significant reduction in GABA levels in the inferior occipital gyrus (IOG) in the ITT and PP populations after supplement intervention and follow up, highlighting the effects of GOS on whole-brain chemistry. In the high anxious subgroup, there was a trend level reduction in the dlPFC due to GOS supplementation, and a reduction in the ACC at follow up, two key brain regions associated with emotional regulation and cognitive function, illustrating a sensitivity of the GABA response in high anxious participants. For glutamate, there were trend level reductions in the GOS group at follow up in the dlPFC of the ITT population, and in the ACC and IOG of the PP population. In the High anxious population, GOS reduced glutamate in the ACC at follow up matching the reduction reported in GABA. Analysis of E/I ratio showed an increase in the high anxious GOS group following intervention in the dlPFC, reflecting the decrease in GABA.

Additionally, the gut microbiome analysis found GOS supplementation to increase *Bifidobacterium* compared to placebo following intervention (*p* = .042, CI[−1.56,−0.02]). Following the intervention, GOS led to an increase in *Bifidobacterium* abundance, though the effect was transient. In the ITT population, abundance rose by 84% at endline but declined by 31.8% at follow-up. Similarly, in the PP population, abundance doubled with a 105% increase at endline, only to decrease sharply by 78.4% at follow-up. This is consistent with the known prebiotic effects of GOS. Interestingly, broader microbiome diversity measures did not show significant changes, suggesting that the effects of GOS were specific to *Bifidobacterium* rather than overall community structure.

In terms of tertiary outcomes, GOS supplementation did not significantly affect psychological variables such as social anxiety, depression, or attentional bias. However, a notable improvement was observed in behavioral reaction times in the high-anxious population. Specifically, participants in the GOS group reacted more quickly than those in th e placebo group in both simple and choice reaction time tasks, suggesting a potential cognitive benefit of GOS in individuals with heightened anxiety. Nutritional intake measures showed no significant changes following GOS supplementation, although energy intake was negatively correlated with trait anxiety, consistent with previous literature ^84^ on the relationship between anxiety and eating behaviors.

In summary, while GOS did not influence immediate psychological improvements, the observed neurochemical and microbiome changes indicate that GOS may have subtle but meaningful effects on the gut-brain axis that could translate into longer-term psychological or cognitive benefits. Several recent supplementation trials support the idea that covert biological processes exhibit more rapid modifications than self-assessed psychological indices. For example in a recent well-designed cross over trial of 28 days probiotic supplementation on emotional reactivity in moderately stressed adults, there were no overt improvements in subjective stress reports, yet improvements in covert responses were evident in emotional reactivity during a cognitive task ^85^. The present study mirrors these effects, whereby covert improvements in GABA levels, *Bifidobacterium* growth, and speed of processing in response times were evident in the GOS group following supplement intervention, but not at the subjective self-report level.

Similarly, in another placebo controlled 28 day probiotic intervention trial of high stress, overweight students with sleep difficulties, covert measures showed a response to intervention (increase in peripheral GABA levels, suppression of HPA axis hormones) while overt self-reported mood did not ^86^. In contrast, a recent placebo-controlled probiotic study revealed both covert and overt effects after six weeks of supplementation. Covert measures, including functional near-infrared spectroscopy (fNIRS) to assess brain activity during a mental arithmetic task and metabolomic analysis of stool samples, as well as overt self-reported improvements in mood, were observed. ^87^. This suggests that behavioural improvements in mood and well-being may take additional time to develop, following the progression of underlying biological and physiological changes.

At the brain level, it was found that GABA reduced due to GOS supplementation. Predominantly, this was seen in the IOG, but also indicated in the dlPFC and ACC in high anxious participants. Our focus on GABA is due to the strong associations with *Bifidobacterium* in the gut microbiome, but we also measured glutamate and the ratio of glutamate to GABA as a metric of cortical excitability, a critical dimension in the neurobiological basis of anxiety. There were trends of lowered glutamate levels via GOS at follow up in the ACC and dlPFC, but not immediately following intervention. An increase in E/I ratio was observed in the dlPFC following supplement intervention for the high anxious group. Overall, the findings indicate that GABA levels decreased in the IOG due to GOS supplementation, with specific effects in the ACC and dlPFC of the high-anxious population. These results suggest a rebalancing of the E/I ratio, potentially reflecting improved cortical responsiveness.

Improved cortical responsiveness in the high anxious population represents a modification in the neural circuitry underpinning anxiety. The reductions in GABA levels in key emotion regulation regions were antithetical to our predictions and yet evident in the high anxious population. Emerging evidence suggests that reductions in ^1^H-MRS measured GABA levels are likely beneficial in anxious states ^36,43,88–90^, despite past inconsistencies in the relationship of prefrontal GABA and anxiety where both positive ^91^ and negative correlations have been found ^92–94^.

Building on the neural over-inhibition hypothesis of anxiety ^36^, which posits that anxious states are sustained by elevated GABA relative to glutamate in the brain, we propose that lowered GABA levels in the GOS group reflect improvements in cortical excitability by rebalancing glutamate levels. For high anxious individuals, increased levels of GABA signal imbalanced cortical networks whereby inhibitory processes associated with GABA levels work overtime on monitoring sensory input and impair higher order cognition ^43^. Here we show that GOS reduces GABA in critical emotion processing regions, bringing the neural circuitry underlying emotional processing into balance. This evidence aligns with neurocomputational accounts of brain networks (e.g., ^43^) that consider neurochemistry critical in coordinating brain function underpinning behaviour.

The IOG is an occipital region of the brain, primarily engaged in visual processing and for that reason functional specialisation and structural maturation are realised earlier than other later developing regions on the brain. It is assumed that due to a state of stable regulation, E/I levels, or GABA and glutamate measured in the IOG would serve as a control metric against the dlPFC and ACC regions in the prefrontal cortex that reach maturity later. However, it is clear from these data that GABA levels are reducing due to GOS. A reduction in GABA in the IOG may signal a path to altered sensory processing, either increasing efficiency, or tuning down responsiveness in visual information processing ^95^. This suggests modifications to brain GABA levels that are not specific only in anxiety, but also generally.

On GOS effects of the gut microbiome, there were clear increases in *Bifidobacterium* following supplement intervention, and marked decreases at follow up. While this is confirmatory of the prebiotic effects of GOS ^96^, and noting a transient pattern, there was no other changes in the gut microbiome composition due to GOS. The selective increase in *Bifidobacterium* abundance is significant for multiple reasons: *Bifidobacterium* are critical for maintaining gut health ^7^, in immunological function ^98^ via production of short chain fatty acids ^99,100^, and in early neurodevelopment ^101^ and beyond ^96^. Coupled with clear changes in neurochemistry measured in vivo, we determined that positive modulation of the gut microbiome with GOS impacts neural circuitry underpinning brain function. While deterministic intervention studies in humans to modulate the gut microbiome and influence brain function have previously used functional and structural neuroimaging of the brain ^102–104^, this is the first to examine neurochemistry directly. These complements supporting evidence in preclinical models. For example, a *B. dentium* intervention has been shown to increase GABA levels in the gut and influence tyrosine in the brain ^105^, the prebiotics GOS and FOS result in altered GABA and 5-HT in the brain ^106^, and probiotic supplementation in disease downregulates GABA ^107^.

This study has several strengths. That it is a randomised double-blind placebo-controlled trial allows deterministic, robust, and unbiased inference on the effects of GOS. Muli-levels of assessment allow us to examine responses along the entire gut brain axis and is the first to measure neurochemical responses in this scenario in vivo, allowing direct observation of critical neurotransmitters such as GABA. The single sex sample enhances study rigor as research indicates that GABA levels in the prefrontal cortex (PFC) differ by sex ^108,109^, and that male levels of GABA and associated inhibitory thresholds in emotion-related outcomes differ in comparison to females ^110,111^. However, the study may be improved by extending the supplement intervention duration to amplify effects at the mental health level, or an alternative approach would be to focus on high anxious participants. No effects on depression or social anxiety were found despite evidence from other studies (e.g., ^112,113^). We did however find improvements in cognitive function, at the level of speed of information processing in reaction rates. This was most prominent in high anxious populations and may be explained by a reduction in anxious states, facilitated by GOS to allow more efficient information processing ^114^.

## Conclusion

While GOS supplementation did not yield significant trait anxiety reductions following intervention, the observed neurochemical changes, gut microbiota modulation, and reaction time improvements in high-anxious individuals indicate GOS’s potential as a modulator of the gut-brain axis. These findings suggest further investigation into longer-term and population-specific effects, especially among individuals with heightened anxiety or altered neurochemical states.

## Acknowledgements

We thank Melissa Basso, Eloise Crowson, Nathalie Dusart, Hannah Eiseman, Chloe Hambleton, Daniella Louise Jones, Paul Knytl, Mariia Malova, and Greta Tomasi for supporting the production of research materials and data in this study.

## Data availability

The data that support the findings of this study are openly available in osf at https://osf.io/bejkn/?view_only=5abce026fb474bd8b6e8a42c94dc52fa

## Trial Registration

The protocol for this study was prospectively registered on https://clinicaltrials.gov/ [NCT03835468] on February 2, 2019.

## CRediT Authorship

**Nicola Johnstone**: conceptualisation, data curation (lead), formal analysis (lead), funding acquisition, investigation (equal), methodology (equal), project administration (equal), visualisation (lead), writing – original draft preparation (lead), writing – review and editing (equal). **Kathrin Cohen Kadosh**: conceptualisation (lead), funding acquisition (lead), investigation (equal), methodology (equal), project administration (equal), writing – review and editing (equal).

## References.

1. Goodman, A., Joyce, R. & Smith, J. P. The long shadow cast by childhood physical and mental problems on adult life. Proc. Natl. Acad. Sci. 108, 6032–6037 (2011).

2. Kieling, C. et al. Child and adolescent mental health worldwide: evidence for action. The Lancet 378, 1515–1525 (2011).

3. Riccio, P. & Rossano, R. The human gut microbiota is neither an organ nor a commensal. FEBS Lett. 594, 3262–3271 (2020).

4. Roswall, J. et al. Developmental trajectory of the healthy human gut microbiota during the first 5 years of life. Cell Host Microbe 29, 765–776 (2021).

5. Valdes, A. M., Walter, J., Segal, E. & Spector, T. D. Role of the gut microbiota in nutrition and health. BMJ 361, k2179 (2018).

6. Leeming, E. R., Johnson, A. J., Spector, T. D. & Roy, C. I. L. Effect of Diet on the Gut Microbiota: Rethinking Intervention Duration. Nutrients 11, 2862 (2019).

7. Gibson, G. R. & Roberfroid, M. B. Dietary Modulation of the Human Colonic Microbiota: Introducing the Concept of Prebiotics. J. Nutr. 125, 1401–1412 (1995).

8. Gibson, G. R., Probert, H. M., Loo, J. V., Rastall, R. A. & Roberfroid, M. B. Dietary modulation of the human colonic microbiota: updating the concept of prebiotics. Nutr. Res. Rev. 17, 259–275 (2004).

9. Gibson, G. R. et al. Expert consensus document: The International Scientific Association for Probiotics and Prebiotics (ISAPP) consensus statement on the definition and scope of prebiotics. Nat. Rev. Gastroenterol. Hepatol. 2017 148 14, 491–502 (2017).

10. Johnstone, N. et al. Anxiolytic effects of a galacto-oligosaccharides prebiotic in healthy females (18–25 years) with corresponding changes in gut bacterial composition. Sci. Rep. 11, 8302 (2021).

11. Schmidt, K. et al. Prebiotic intake reduces the waking cortisol response and alters emotional bias in healthy volunteers. Psychopharmacology (Berl.) 232, 1793–1801 (2015).

12. Desbonnet, L. et al. Gut microbiota depletion from early adolescence in mice: Implications for brain and behaviour. Brain. Behav. Immun. 48, 165–173 (2015).

13. Murray, E. et al. Probiotic consumption during puberty mitigates LPS-induced immune responses and protects against stress-induced depression- and anxiety-like behaviors in adulthood in a sex-specific manner. Brain. Behav. Immun. 81, 198–212 (2019).

14. Provensi, G. et al. Preventing adolescent stress-induced cognitive and microbiome changes by diet. Proc. Natl. Acad. Sci. 116, 9644–9651 (2019).

15. Wu, J. et al. Changes in gut viral and bacterial species correlate with altered 1,2-diacylglyceride levels and structure in the prefrontal cortex in a depression-like non-human primate model. Transl. Psychiatry 12, (2022).

16. Diaz Heijtz, R. Fetal, neonatal, and infant microbiome: Perturbations and subsequent effects on brain development and behavior. Semin. Fetal. Neonatal Med. 21, 410–417 (2016).

17. Heijtz, R. D. et al. Normal gut microbiota modulates brain development and behavior. Proc. Natl. Acad. Sci. U. S. A. 108, 3047–3052 (2011).

18. Nicholson, J. K. et al. Host-Gut Microbiota Metabolic Interactions. Proc Natl Acad Sci USA 5, 45– 54 (2011).

19. Long-Smith, C. et al. Microbiota-Gut-Brain Axis: New Therapeutic Opportunities. Annu. Rev. Pharmacol. Toxicol. 60, 477–502 (2020).

20. Borre, Y. E., Moloney, R. D., Clarke, G., Dinan, T. G. & Cryan, J. F. The Impact of Microbiota on Brain and Behavior: Mechanisms & Therapeutic Potential. in Microbial Endocrinology: The Microbiota-Gut-Brain Axis in Health and Disease (eds. Lyte, M. & Cryan, J. F.) 373–403 (Springer New York, New York, NY, 2014). doi:10.1007/978-1-4939-0897-4_17.

21. Luczynski, P. et al. Growing up in a Bubble: Using Germ-Free Animals to Assess the Influence of the Gut Microbiota on Brain and Behavior. Int. J. Neuropsychopharmacol. 19, pyw020 (2016).

22. McVey Neufeld, K.-A., Luczynski, P., Seira Oriach, C., Dinan, T. G. & Cryan, J. F. What’s bugging your teen?—The microbiota and adolescent mental health. Neurosci. Biobehav. Rev. 70, 300– 312 (2016).

23. McVey Neufeld, S. F., Ahn, M., Kunze, W. A. & McVey Neufeld, K.-A. Adolescence, the Microbiota-Gut-Brain Axis, and the Emergence of Psychiatric Disorders. Biol. Psychiatry (2023) doi:10.1016/j.biopsych.2023.10.006.

24. O’Mahony, S. M. et al. The enduring effects of early-life stress on the microbiota–gut–brain axis are buffered by dietary supplementation with milk fat globule membrane and a prebiotic blend. Eur. J. Neurosci. 51, 1042–1058 (2020).

25. Barrett, E., Ross, R. P., O’Toole, P. W., Fitzgerald, G. F. & Stanton, C. γ-Aminobutyric acid production by culturable bacteria from the human intestine. J. Appl. Microbiol. 113, 411–417 (2012).

26. Duranti, S. et al. Bifidobacterium adolescentis as a key member of the human gut microbiota in the production of GABA. Sci. Rep. 10, 14112 (2020).

27. Braga, J. D., Thongngam, M. & Kumrungsee, T. Gamma-aminobutyric acid as a potential postbiotic mediator in the gut–brain axis. Npj Sci. Food 8, 16 (2024).

28. Mann, E. R., Lam, Y. K. & Uhlig, H. H. Short-chain fatty acids: linking diet, the microbiome and immunity. Nat. Rev. Immunol. 24, 577–595 (2024).

29. Silva, Y. P., Bernardi, A. & Frozza, R. L. The Role of Short-Chain Fatty Acids From Gut Microbiota in Gut-Brain Communication. Front. Endocrinol. 11, (2020).

30. Bercik, P., Collins, S. M. & Verdu, E. F. Microbes and the gut-brain axis. Neurogastroenterol. Motil. 24, 405–413 (2012).

31. Carabotti, M., Scirocco, A., Maselli, M. A. & Severi, C. The gut-brain axis: interactions between enteric microbiota, central and enteric nervous systems. Ann. Gastroenterol. Q. Publ. Hell. Soc. Gastroenterol. 28, 203 (2015).

32. Ben-Ari, Y. Excitatory actions of gaba during development: the nature of the nurture. Nat. Rev. Neurosci. 3, 728–739 (2002).

33. Owens, D. F. & Kriegstein, A. R. Is there more to GABA than synaptic inhibition? Nat Rev Neurosci 3, 715–727 (2002).

34. Thomson, A. R. et al. The developmental trajectory of 1H-MRS brain metabolites from childhood to adulthood. Cereb. Cortex 34, bhae046 (2024).

35. Porges, E. C., Jensen, G., Foster, B., Edden, R. A. E. & Puts, N. A. J. The trajectory of cortical gaba across the lifespan, an individual participant data meta-analysis of edited mrs studies. eLife 10, (2021).

36. Johnstone, N. & Cohen Kadosh, K. Excitatory and inhibitory neurochemical markers of anxiety in young females. Dev. Cogn. Neurosci. 66, 101363 (2024).

37. Larsen, B. et al. A developmental reduction of the excitation:inhibition ratio in association cortex during adolescence. Sci. Adv. 8, eabj8750 (2022).

38. Cohen Kadosh, K., Krause, B., King, A. J., Near, J. & Cohen Kadosh, R. Linking GABA and glutamate levels to cognitive skill acquisition during development. Hum Brain Mapp 36, 4334– 4345 (2015).

39. McKeon, S. D. et al. Prefrontal excitation/inhibition balance supports adolescent enhancements in circuit signal to noise ratio. Prog. Neurobiol. 243, 102695 (2024).

40. Parr, A. C., Sydnor, V. J., Calabro, F. J. & Luna, B. Adolescent-to-adult gains in cognitive flexibility are adaptively supported by reward sensitivity, exploration, and neural variability. Curr. Opin. Behav. Sci. 58, 101399 (2024).

41. Uhlhaas, P. J. et al. Towards a youth mental health paradigm: a perspective and roadmap. Mol. Psychiatry 1–11 (2023) doi:10.1038/s41380-023-02202-z.

42. Khona, M. & Fiete, I. R. Attractor and integrator networks in the brain. Nat. Rev. Neurosci. 23, 744–766 (2022).

43. LeDuke, D. O., Borio, M., Miranda, R. & Tye, K. M. Anxiety and depression: A top-down, bottom-up model of circuit function. Ann. N. Y. Acad. Sci. 1525, 70–87 (2023).

44. Johnstone, N. et al. Nutrient Intake and Gut Microbial Genera Changes after a 4-Week Placebo Controlled Galacto-Oligosaccharides Intervention in Young Females. Nutrients 13, 4384 (2021).

45. Etkin, A., Büchel, C. & Gross, J. J. The neural bases of emotion regulation. Nat Rev Neurosci 16, 693–700 (2015).

46. Spielberger, C. D., Gorsuch, R. L., Lushene, R., Vagg, P. R. & Jacobs, G. A. State-Trait Anxiety Inventory for Adults. (Mind Garden Inc., Palo Alto, California, 1983).

47. La Greca, A. M. Manual for the Social Anxiety Scales for Children and Adolescents - Revised. (University of Miami, Miami, Florida, 1999).

48. Beck, A. T., Steer, R. A. & Brown, G. K. BDI-II: Beck Depression Inventory Manual. (Psychological Corporation, San Antonio, Texas, 1996).

49. MacLeod, C., Mathews, A. & Tata, P. Attentional bias in emotional disorders. J. Abnorm. Psychol. 95, 15–20 (1986).

50. Deary, I. J., Liewald, D. & Nissan, J. A free, easy-to-use, computer-based simple and four-choice reaction time programme: The Deary-Liewald reaction time task. Behav. Res. Methods 43, 258– 268 (2011).

51. Qualtrics. Qualtircs (2005).

52. Buysse, D. J., Reynolds, C. F., Monk, T. H., Berman, S. R. & Kupfer, D. J. The Pittsburgh Sleep Quality Index: a new instrument for psychiatric practice and research. Psychiatry Res. 28, 193– 213 (1989).

53. Steiner, M. et al. The premenstrual symptoms screening tool revised for adolescents (PSST-A): Prevalence of severe PMS and premenstrual dysphoric disorder in adolescents. Arch. Womens Ment. Health 14, 77–81 (2011).

54. Craig, C. L. et al. International physical activity questionnaire: 12-Country reliability and validity. Med. Sci. Sports Exerc. 35, 1381–1395 (2003).

55. Nutritics. (2019).

56. Peirce, J., Hirst, R. & MacAskill, M. Building Experiments in PsychoPy. (SAGE, London, 2022).

57. Hillmann, B., et al. Evaluating the Information Content of Shallow Shotgun Metagenomics. mSystems 3, 10.1128/msystems.00069-18 (2018).

58. Chen, S., Zhou, Y., Chen, Y. & Gu, J. fastp: an ultra-fast all-in-one FASTQ preprocessor. Bioinformatics 34, i884–i890 (2018).

59. Breiman, L. Random forests. Mach. Learn. 45, 5–32 (2001).

60. Buchfink, B., Xie, C. & Huson, D. H. Fast and sensitive protein alignment using DIAMOND. Nat. Methods 12, 59–60 (2015).

61. Cantarel, B. et al. The Carbohydrate-Active Enzymes database (CAZy): an expert resource for Glycogenomics. Nucleic Acids Res. 37, D233–D238 (2009).

62. Feng, Q. et al. Gut microbiome development along the colorectal adenoma-carcinoma sequence. Nat. Commun. 6, 6528 (2015).

63. Huson, D. H., Mitra, S., Ruscheweyh, H.-J., Weber, N. & Schuster, S. C. Integrative analysis of environmental sequences using MEGAN4. Genome Res. 21, 1552–1560 (2011).

64. Kanehisa, M., Goto, S., Furumichi, M., Tanabe, M. & Hirakawa, M. KEGG for representation and analysis of molecular networks involving diseases and drugs. Nucleic Acids Res. 38, D355–D360 (2006).

65. Karlsson, F. H. et al. Gut metagenome in European women with normal, impaired and diabetic glucose control. Nature 498, 99–103 (2013).

66. Langmead, B. & Salzberg, S. L. Fast gapped-read alignment with Bowtie 2. Nat. Methods 9, 357– 359 (2012).

67. Li, D., Liu, C.-M., Luo, R., Sadakane, K. & Lam, T.-W. MEGAHIT: an ultra-fast single-node solution for large and complex metagenomics assembly via succinct de Bruijn graph. Bioinformatics 31, 1674–1676 (2015).

68. Li, W. & Godzik, A. Cd-hit: a fast program for clustering and comparing large sets of protein or nucleotide sequences. Bioinformatics 22, 1658–1659 (2006).

69. Qin, J. et al. A metagenome-wide association study of gut microbiota in type 2 diabetes. Nature 490, 55–60 (2010).

70. Karlsson, F. H. et al. Symptomatic atherosclerosis is associated with an altered gut metagenome. Nat. Commun. 3, 1245 (2012).

71. Scher, J. U. et al. Expansion of intestinal Prevotella copri correlates with enhanced susceptibility to arthritis. eLife 2, e01202 (2013).

72. Qin, J. et al. A human gut microbial gene catalogue established by metagenomic sequencing. Nature 464, 59–65 (2010).

73. Zhu, W., Lomsadze, A. & Borodovsky, M. Ab initio gene identification in metagenomic sequences. Nucleic Acids Res. 38, e132 (2010).

74. Li, J. et al. An integrated catalog of reference genes in the human gut microbiome. Nat. Biotechnol. 32, 834–841 (2014).

75. Mallick, H. et al. Multivariable association discovery in population-scale meta-omics studies. PLOS Comput. Biol. 17, e1009442 (2021).

76. Lin, H. & Peddada, S. D. Multigroup analysis of compositions of microbiomes with covariate adjustments and repeated measures. Nat. Methods 21, 83–91 (2024).

77. Mlynárik, V., Gambarota, G., Frenkel, H. & Gruetter, R. Localized short-echo-time proton MR spectroscopy with full signal-intensity acquisition. Magn. Reson. Med. 56, 965–970 (2006).

78. Simpson, R., Devenyi, G. A., Jezzard, P., Hennessy, T. J. & Near, J. Advanced processing and simulation of MRS data using the FID appliance (FID-A)—An open source, MATLAB-based toolkit. Magn. Reson. Med. 77, 23–33 (2017).

79. Harris, A. D., Puts, N. A. J. & Edden, R. A. E. Tissue correction for GABA-edited MRS: Considerations of voxel composition, tissue segmentation, and tissue relaxations. J. Magn. Reson. Imaging JMRI 42, 1431–1440 (2015).

80. Edden, R. A. E., Puts, N. A. J., Harris, A. D., Barker, P. B. & Evans, C. J. Gannet: A batch-processing tool for the quantitative analysis of gamma-aminobutyric acid-edited MR spectroscopy spectra. J. Magn. Reson. Imaging 40, 1445–1452 (2014).

81. Ashburner, J. & Friston, K. J. Unified segmentation. NeuroImage 26, 839–851 (2005).

82. Provencher, S. W. Estimation of metabolite concentrations from localizedin vivo proton NMR spectra. Magn. Reson. Med. 30, 672–679 (1993).

83. R Core Team. R: A Language and environment for statistical computing. R Foundation for Statistical Computing. (2023).

84. Keck, M. M. et al. Examining the Role of Anxiety and Depression in Dietary Choices among College Students. Nutrients 12, 2061 (2020).

85. Casertano, M. et al. Gaba-producing lactobacilli boost cognitive reactivity to negative mood without improving cognitive performance: A human Double-Blind Placebo-Controlled Cross-Over study. Brain. Behav. Immun. 122, 256–265 (2024).

86. Li, J. et al. Effects of Bifidobacterium breve 207-1 on regulating lifestyle behaviors and mental wellness in healthy adults based on the microbiome-gut-brain axis: a randomized, double-blind, placebo-controlled trial. Eur. J. Nutr. 63, 2567–2585 (2024).

87. Mutoh, N. et al. Bifidobacterium breve M-16V regulates the autonomic nervous system via the intestinal environment: A double-blind, placebo-controlled study. Behav. Brain Res. 460, 114820 (2024).

88. Page, C. E. & Coutellier, L. Prefrontal excitatory/inhibitory balance in stress and emotional disorders: Evidence for over-inhibition. Neurosci. Biobehav. Rev. 105, 39–51 (2019).

89. Perica, M. I. et al. Development of frontal GABA and glutamate supports excitation/inhibition balance from adolescence into adulthood. Prog. Neurobiol. 219, 102370 (2022).

90. McKeon, S. D. et al. Aperiodic EEG and 7T MRSI evidence for maturation of E/I balance supporting the development of working memory through adolescence. 2023.09.06.556453 Preprint at 10.1101/2023.09.06.556453 (2023).

91. Delli Pizzi, S. GABA content within the ventromedial prefrontal cortex is related to trait anxiety. Soc Cogn Affect Neurosci 11, 758–766 (2016).

92. Hasler, G. Prefrontal Cortical Gamma-Aminobutyric Acid Levels in Panic Disorder Determined by Proton Magnetic Resonance Spectroscopy. Biol Psychiatry 65, 273–275 (2009).

93. Hasler, G. Association between prefrontal glutamine levels and neuroticism determined using proton magnetic resonance spectroscopy. Transl Psychiatry 9, 1–8 (2019).

94. Long, Z. Decreased GABA levels in anterior cingulate cortex/medial prefrontal cortex in panic disorder. Prog Neuropsychopharmacol Biol Psychiatry 44, 131–135 (2013).

95. Umesawa, Y., Atsumi, T., Chakrabarty, M., Fukatsu, R. & Ide, M. GABA Concentration in the Left Ventral Premotor Cortex Associates With Sensory Hyper-Responsiveness in Autism Spectrum Disorders Without Intellectual Disability. Front. Neurosci. 14, 482 (2020).

96. Cryan, J. F. & Dinan, T. G. Mind-altering microorganisms: the impact of the gut microbiota on brain and behaviour. Nat. Rev. Neurosci. 13, 701–712 (2012).

97. Biavati, B. & Mattarelli, P. Bifidobacterium ruminantium sp. nov. and Bifidobacterium merycicum sp. nov. from the Rumens of Cattle. Int. J. Syst. Evol. Microbiol. 41, 163–168 (1991).

98. Sivan, A. et al. Commensal Bifidobacterium promotes antitumor immunity and facilitates anti-PD-L1 efficacy. Science 350, 1084–1089 (2015).

99. Fukuda, S. et al. Bifidobacteria can protect from enteropathogenic infection through production of acetate. Nature 469, 543–549 (2011).

100. Maslowski, K. M. et al. Regulation of inflammatory responses by gut microbiota and chemoattractant receptor GPR43. Nature 461, 1282–1286 (2009).

101. Sudo, N. et al. Postnatal microbial colonization programs the hypothalamic-pituitary-adrenal system for stress response in mice. J. Physiol. 558, 263–275 (2004).

102. Pinto-Sanchez, M. I. et al. Probiotic Bifidobacterium longum NCC3001 Reduces Depression Scores and Alters Brain Activity: A Pilot Study in Patients With Irritable Bowel Syndrome. Gastroenterology 153, 448–459.e8 (2017).

103. Tillisch, K. et al. Consumption of Fermented Milk Product With Probiotic Modulates Brain Activity. Gastroenterology 144, 1394–1401.e4 (2013).

104. Tillisch, K. et al. Brain structure and response to emotional stimuli as related to gut microbial profiles in healthy women. Psychosom. Med. 79, 905 (2017).

105. Luck, B. et al. Neurotransmitter profiles are altered in the gut and brain of mice mono-associated with Bifidobacterium dentium. Biomolecules 11, (2021).

106. Zhang, S. et al. Prebiotics modulate the microbiota–gut–brain axis and ameliorate cognitive impairment in APP/PS1 mice. Eur. J. Nutr. 62, 2991–3007 (2023).

107. Zhao, H. et al. Ingestion of Lacticaseibacillus rhamnosus Fmb14 prevents depression-like behavior and brain neural activity via the microbiota-gut-brain axis in colitis mice. Food Funct. 14, 1909–1928 (2023).

108. Beck, D. et al. Puberty differentially predicts brain maturation in male and female youth: A longitudinal ABCD Study. Dev. Cogn. Neurosci. 61, 101261 (2023).

109. Juraska, J. M. & Drzewiecki, C. M. Cortical reorganization during adolescence: What the rat can tell us about the cellular basis. Dev. Cogn. Neurosci. 45, 100857 (2020).

110. Cohen, J. E. et al. Neural response to stress differs by sex in young adulthood. Psychiatry Res. Neuroimaging 332, 111646 (2023).

111. Strasser, A., Xin, L., Gruetter, R. & Sandi, C. Nucleus accumbens neurochemistry in human anxiety: A 7 T 1H-MRS study. Eur Neuropsychopharmacol 29, 365–375 (2019).

112. Methiwala, H. N. et al. Gut microbiota in mental health and depression: Role of pre/pro/synbiotics in their modulation. Food Funct. 12, 4284–4314 (2021).

113. Ritz, N. L., et al. Social anxiety disorder-associated gut microbiota increases social fear. Proc. Natl. Acad. Sci. 121, e2308706120 (2024).

114. Bloemendaal, M. et al. Probiotics-induced changes in gut microbial composition and its effects on cognitive performance after stress: exploratory analyses. Transl. Psychiatry 11, 300 (2021).

